# A population model of age and gender specific cardiac troponin levels

**DOI:** 10.1101/2024.12.20.24319414

**Authors:** Alexandru Dan Corlan

## Abstract

The range of the high-sensitivity cardiac troponin (HSCTN) values in the general population increases progressively with age More accurate normal limits for HSCTN, taking into account age and gender, are needed for the differential diagnosis and evaluation of the prognostic significance of increases that do not reach the vendor-supplied upper reference limits (URL). We performed an analysis of the high sensitivity cardiac troponin (HSCTN) of 21743 individuals, representative for the general US population, that were studied in the NHANES survey performed by the Center for Disease Control of the USA. The vendor supplied URL values are typically several times higher than the actual upper limit in subjects under 40 and correspond to the URL at ages between 51 and 84 for specific vendors and genders. For each HSCTN test variant, we considered each one year age group between 1 and 85 and either gender. The distribution of the logarithmed HSCTN for a given test variant and gender, in each subsample i, is relatively close to Gaussian 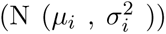. Two quadratic models, for the 1-15 and the 16-85 age ranges, were found to fit well with the *µ_i_*, while the *σ_i_* follow a linear model. We propose a standardised HSCTN based on such theoretical distributions, that are independent of testing kit, age and gender.

## 1 Introduction

A sudden increase in the high sensitivity cardiac troponin (HSCTN) level, above the upper reference limit (URL), is an early, high specificity indicator for an acute myocardial infarction (AMI). It is currently the main indicator used for early myocardial infarction diagnosis [1, 4, 13].

At the same time, there is a growing body of evidence for the prognostic and diagnostic value of relatively high HSCTN levels that may be associated to non-AMI acute coronary syndromes [1, 4] and other cardiac and extracardiac conditions such as heart failure [8], hypertrophy[8, 2, 10], cardiomyopathy [7], arterial hypertension [10] or kidney disease [11], as well as for general mortality [9].

The URL is a level established by the producer of a HSCTN test kit, that corresponds to the 99-th centile in the general population. Diagnostic and prognostic meaning of a HSCTN test revolves around this level. There are usually two different gender specific URLs. They are determined by estimating the 99-th centile in HSCTN measurements from samples of the male and female adult population, typically around 500 cases.

Recently, a strong relation between the HSCTN and age was reported, both for adults [3] and for children [6]. See [6] for a discussion of how puberty influences HSCTN. In older adults, after the age of 40, the HSCTN values increase with age. Thus, in a sample of the general population, the 99-th centile will reflect the HSCTN of the older individuals. This does not raise a significant issue for the AMI diagnostic, especially in a type 1 MI, when the HSCTN increase is both acute and much higher than the URL, irrespective of age. However, it could theoretically make the detection of other, smaller, possibly chronic HSCTN increases less sensitive in younger patients.

There is increasing awareness of the need for age and gender URLs [3, 12] and also studies that differentiate the URL in older subjects vs younger subjects [5, 14]

The aim of this study is to establish a model for the calculation of the URL of a specific HSCTN test as a continuous function of age and gender, at any age, and a method to compute a test-kit, age and gender independent indicator, based on HSCTN.

## 2 Methods

We used a publicly available dataset in 21743 individuals, aged 1 to 85, that can be found at https://wwwn.cdc.gov/nchs/nhanes and was reported in [9]. Blood samples of participants in a general health survey in the United States were stored, between 1999 and 2004. Twenty years later, the samples were thawed and a variety of laboratory tests were performed on them, in particular four HSCTN tests: a Roche test for troponin T, and three tests for troponin I (Abbott, Siemens and Ortho). All results are available online at the above address.

We extracted these data from these files (DEMO.XPT, DEMO_B.XPT, DEMO_C.XPT, SSTROP_A.XPT) and joined them by the SEQN field. The vendor specific URL was taken from [9]. We call the resulting dataset, below, “the NHANES dataset”.

The stochastic model we built for describing the population is as follows. We consider groups by sex (*s*) and one year age group (*a*). For each group, four random variables are modelled (as dependent variables), representing the high sensitivity cardiac troponins T with the Roche method (TR), troponin I with the Abbott (IA), Siemens (IS) and Ortho(IO) method respectively. When we refer to either method, we denote that by *Y_M_* where M could be any of the methods TR, IA, IS, IO.

For each group, the logarithm of any of the troponin results, ln *Y_M,a,s_*, has a Gaussian distribution with mean *µ_M,a,s_* and standard deviation *σ_M,a,s_*.

For each dependent variable, gender and age, *µ* and *σ* are computed using the coefficients *α_σ_, β_σ_, α_µ_, β_µ_, γ_µ_*. There are two sets of coefficients for *µ*, one for prepuberal children (until 15), marked below with a (’), and one for older individuals (16 and above).

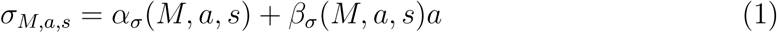

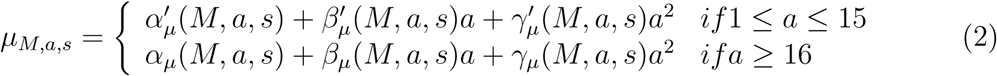

These coefficients, for each age, gender and method, were estimated through quadratic and linear fitting, as shown in the results section.

We use sample medians as data for estimating the *µ_M,a,s_*, rather than sample means, because each age/gender sample probably contains a large proportion of normals, the logarithm of which we presume to be normally distributied, and a small proportion of subjects with various conditions, that appear as outliers. As the median is more robust to the efect of the outliers compared to the sample mean, and is also an estimate of *µ* in the Gaussian distribution, we choose the median.

The cumulative probability distribution for a given HSCTN value, *y*, is computed as:

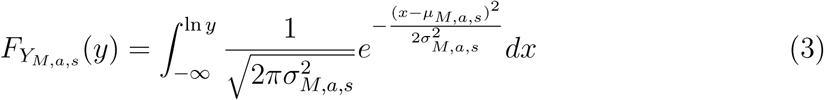

We find the theoretical URL for a given percentile, *c*, by solving equation 4 for URL.

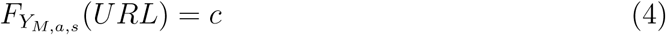

## 3 Results

Distributions of method, gender and age subgroups in the NHANES dataset are shown in figure 1, with larger versions in supplementary figures S1–S4. It can be observed that the distributions are asymmetrical. The distributions of the logarithms of the measurements are shown in figure 2a supplementary figures S5–S8.

**Figure 1:**
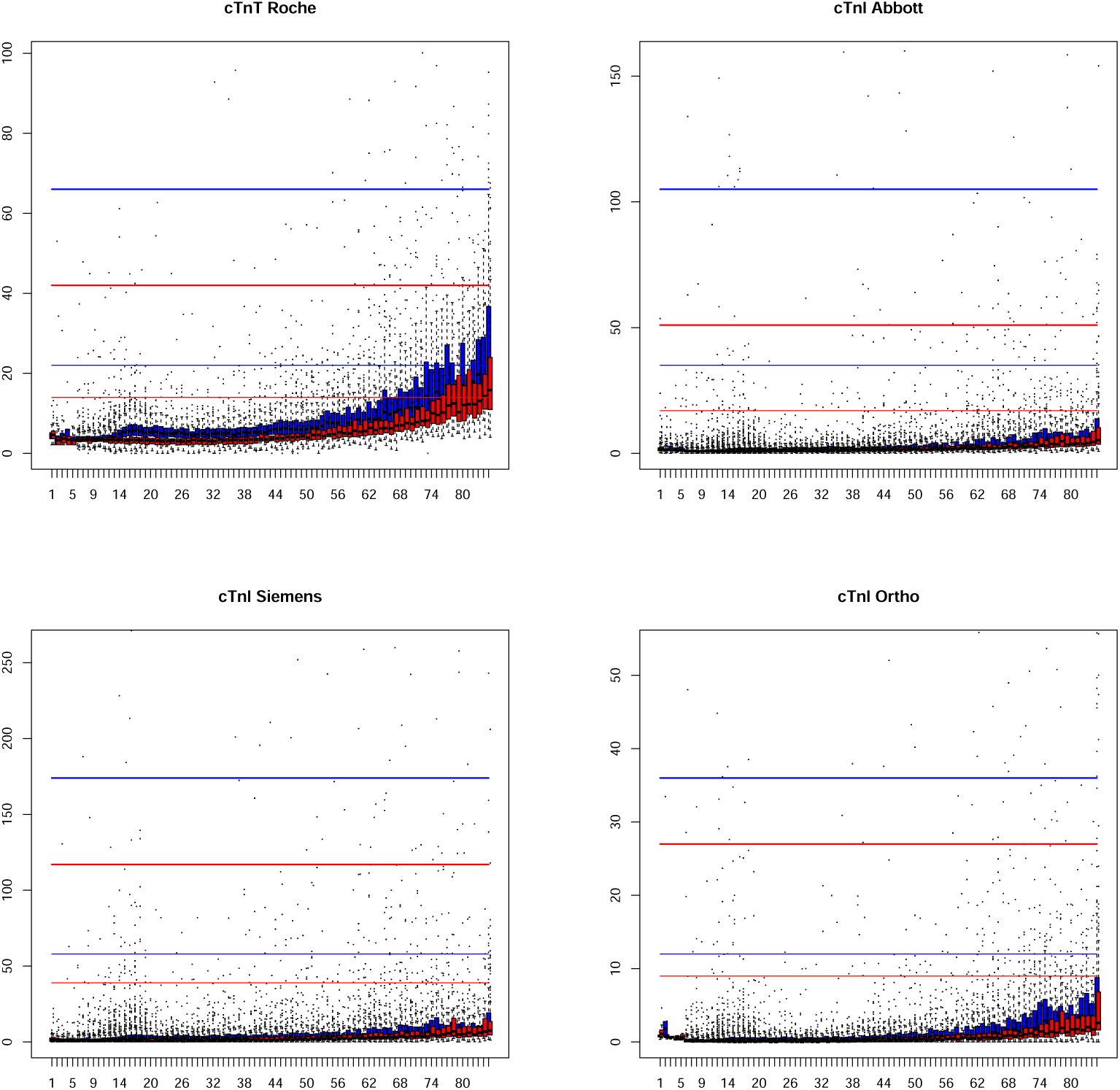
HSCTN distribution by age and gender. Red: females. Blue: males. Horizontal axis: age. Vertical axis: Troponin in ng/L. Boxes: median and first and third quartile. Thin horizontal lines: vendor-supplied upper reference level (URL). Thick horizontal lines: 3 times the URL. The differences between the URL and the actual, age depedent, range of values in the general population can be observed.

**Figure 2:**
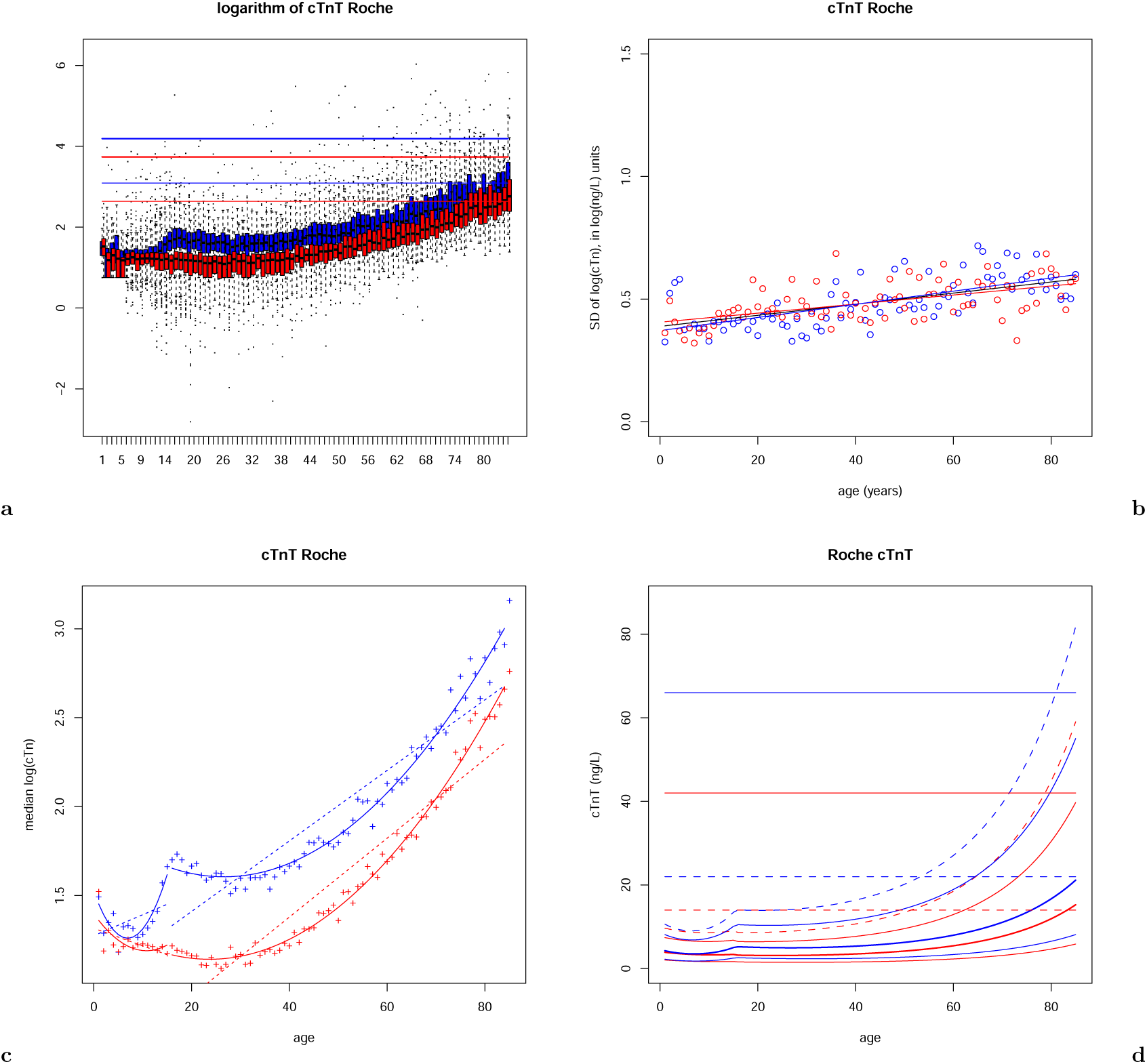
Models of the Roche cTnT distribution. In all pannels—red: females, blue: males, thin horizontal lines: upper limit established by manufacturer, thick horizontal lines: 3 times the upper limit established by manufacturer. a) Logarithm of cTnT Roche, by age and gender; boxes: median and first and third quartile. b) Standard deviations of logarithms of cTnT Roche distributions by age and gender. Line: linear regression of standard deviations as a function of age, for each gender, black for both males and females. c) Medians (crosses) of logarithms of cTnI Abbott distributions by age and gender. Dashed lines: linear regression of medians as a function of age for each gender. Curves: quadratic regressions of medians as a function of age for each gender, over1–15 (prepuberal children) and 16–85 age ranges. d) Model centile lines of age and gender dependent models for Roche cTnT. Thick curve: median. Thin continuous curves: 0.05 and 0.95 centiles. Dashed curve: 0.99 centile.

The medians of the distributions in each age/gender subgroup, and linear and quadratic regression lines are shown in figure 2b for the Roche troponin T method, and in supplementary figures: S10, S12, S14 and S16 for all methods. We observe that linear regressions (dashed lines) do not fit well, even if different regression lines are used for children and adults. Quadratic regressions, however, fit well. The parameters *α_µ_*(*M, a, s*), *β_µ_*(*M, a, s*), *γ_µ_*(*M, a, s*) for each age range, resulting from these qudratic regressions, are shown in table 1.

**Table 1:**
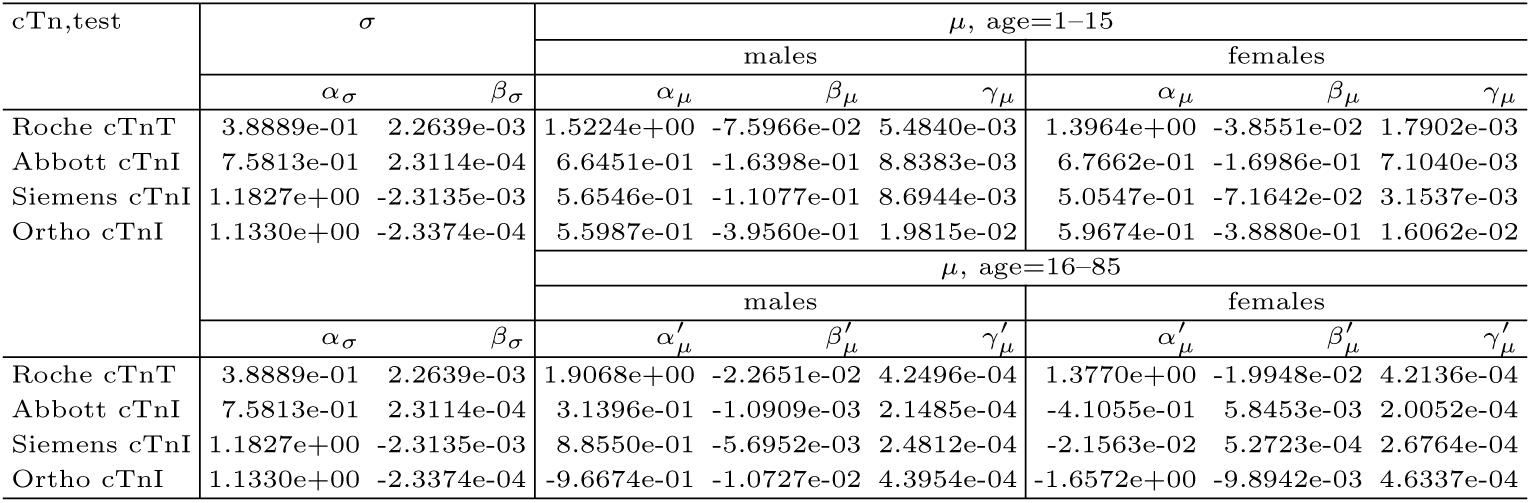
Regression parameters.

The sample standard deviations, as well as the linear regression line of each group are shown in figure 2c and supplementary figures: S9, S11, S13 and S15. We see that a linear regression for the whole age range provides a good fit, and also that the differences between genders are quite small, thus we choose a single linear law for the whole population, for each method.

The coefficients for computing *σ_M,a,s_* for each cTn measurement method and gender are also shown in table 1.

Various quantiles (0.05, 0.5, 0.95, 0.99) of the theoretical distributions *Y_M,a,s_* for each cTn measurement method, by age and gender are shown in figure 2d, for Roche Troponin T, and supplementary figures S17–S20 for all methods, together with their relationship to the gender-specific URL indicated by the manufacturer for each measurement method, and the 3 times URL level that is sometimes used for acute myocardial infarction diagnostic.

The theoretical age and gender-dependent URLs are tabulated in table 2 for the 99-th centile. More detailed tables, also for the other centiles are in the supplementary tables S1, S2 and S3. Table 3 presents the ages where the model-derived 99-th centile URL corresponds to the vendor specified URL.

**Table 2:** General population limits for the 99-th percentile.

| Age | males |  |  |  | females |  |  |  |
| --- | --- | --- | --- | --- | --- | --- | --- | --- |
|  | cTnT | cTnI |  |  | cTnT | cTnI |  |  |
|  | Roche | Abbott | Siemens | Orhto | Roche | Abbott | Siemens | Orhto |
| Limits: | 22 | 35 | 58 | 12 | 14 | 17 | 39 | 9 |
| 1 | 11 | 10 | 25 | 17 | 10 | 10 | 24 | 17 |
| 5 | 9 | 6 | 19 | 6 | 9 | 6 | 19 | 5 |
| 10 | 10 | 5 | 21 | 3 | 9 | 4 | 16 | 3 |
| 15 | 13 | 7 | 34 | 6 | 9 | 4 | 17 | 3 |
| 20 | 14 | 9 | 34 | 5 | 9 | 5 | 15 | 3 |
| 25 | 14 | 9 | 34 | 5 | 9 | 5 | 16 | 3 |
| 30 | 14 | 10 | 34 | 6 | 9 | 6 | 17 | 3 |
| 35 | 15 | 10 | 35 | 6 | 10 | 6 | 18 | 3 |
| 40 | 16 | 11 | 36 | 7 | 11 | 7 | 19 | 4 |
| 45 | 18 | 12 | 38 | 8 | 12 | 8 | 21 | 4 |
| 50 | 20 | 13 | 41 | 9 | 13 | 9 | 23 | 5 |
| 55 | 23 | 15 | 44 | 11 | 16 | 10 | 26 | 6 |
| 60 | 27 | 17 | 48 | 13 | 18 | 12 | 30 | 8 |
| 65 | 32 | 19 | 53 | 16 | 22 | 14 | 35 | 10 |
| 70 | 40 | 22 | 59 | 21 | 28 | 16 | 41 | 12 |
| 75 | 49 | 26 | 67 | 27 | 35 | 19 | 48 | 16 |
| 80 | 63 | 30 | 77 | 36 | 45 | 23 | 58 | 22 |
| 85 | 82 | 36 | 89 | 49 | 59 | 28 | 70 | 31 |

**Table 3:** Age when the model URL equals the vendor supplied URL.

| Test kit | males | females |
| --- | --- | --- |
| Roche cTnT | 53 | 51 |
| Abbott cTnI | 84 | 71 |
| Siemens cTnI | 69 | 68 |
| Ortho cTnI | 57 | 63 |

Figure 3 shows, for each vendor method, plots of each percentile in the 1–99 range and the actual proportion of cases below the respective model centile line. The distance between each point and the identity line indicates the expected error when using the theoretical model to estimate that percentile in the population, or the goodness of fit of the model to the data in the NHANES dataset. It can be seen that the models closely describe the data at each percentile.

**Figure 3:**
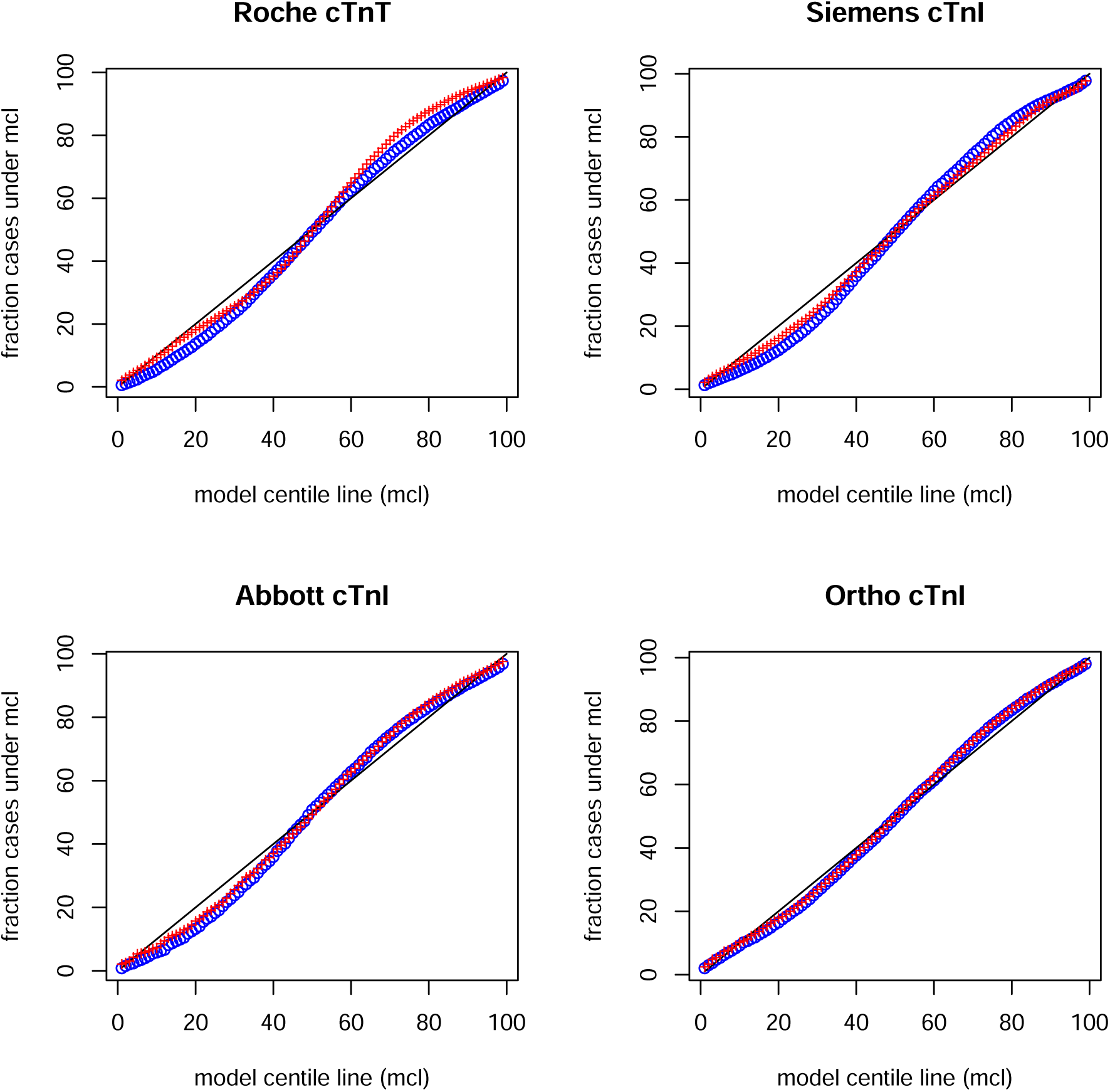
Q-Q plots of the overall model. Red crosses: females. Blue circles: males. The expected centile according to the model is on the horizontal axis. The actual fraction in the whole sample on the vertical axis. It can be seen that each of the 8 models closely follow the distribution of the population.

## 4 Discussion

Our results confirm the results of Franzini et al., [3], that showed a marked increase of the HSCTN values with age, beyond 40 years old, and also the results of Kiess et al., [6] who found that the relatively high HSCTN values in neonates decrease during childhood and then increase during puberty, especially in boys. In our data, early adulthood years are characterised by a relatively stable range of normal values, that starts to increase towards the fourth decade, a process that progresses with every decade of age.

The vendor supplied URL corresponds to that found in the population at ages ranging from 51 to 84 (see supplementary table S1 or table 3), depending on manufacturer and gender. This variability possibly results from the age distribution of the sample used for URL determination.

For younger ages, the vendor supplied URL is 200-300% higher than the actual highest value found in the population. For older ages, abnormal HSCTN values that are above the vendor-supplied URL, sometimes even above 3 times the URL, are found in the general population.

This could result in a significant number of false positive HSCTN diagnoses, especially in older people, and false negative diagnoses in younger people. The fact that a HSCTN increase may result through numerous pathogenic processes complicates the differential diagnostic.

As stated in the introduction, with the exception of the acute phase of many type 1 myocardial infarction cases, where the HSCTN raises, in hours, to values several times above the vendor supplied URL, the use of a single HSCTN URL for all ages may increase the difficulty of the differential diagnosis. This is a problem for various phases of acute coronary syndromes, as described in [13], but also for many other conditions that also lead to relative HSCTN increases.

We proposed a five-parameter population model of the HSCTN, from which more accurate age and gender-specific URL can be computed. For each gender and for each type of testing kit, the five parameters need to be determined experimentally.

Using our models, it is possible to predict the expected 99-th percentile URL at any age. Due to the relatively close fit between observed cumulative frequency and theoretically predictive probability (figure 3), it is also possible to compute, given a HSCTN value, a standardized HSCTN, independent of age, gender, and kit specifics. For levels between 0 and 99, this standardized HSCTN would represent the estimated proportion of the general population that has a lower value than the measured HSCTN. For values above 100, the standardized HSCTN would the percentage above the age and gender specific URL.

Given *L*_99_, the solution of equation 4 for *c* = 0.99, for a measurement *y*, the normalised HSCTN, T, would be, with the notations from equation 3:

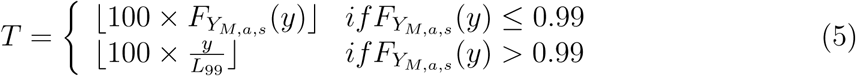

Because the standard, vendor supplied URL has a random relation with the age-dependent URL curve, and the variability of this relation across vendors is high, it is not possible to infer the latter from the former. A new sampling method would be necessary to determine the age-dependent URL curve for each type of testing kit.

Our expectation that the standardized URL, T, would have better diagnostic and prognostic power than the HSCTN and its relationship with the vendor-supplied URL, while intuitive, needs to be experimentally verified. Nevertheless, this verification can be performed, to a significant extent, in existing datasets.

In summary, we proposed a model of the testing kit, gender and age-dependent HSCTN in the general population that closely describes the probability distribution of these values and results in more accurate upper reference limits. This model could become the basis of standardization of the HSCTN measurements.

## Data Availability

All data produced in the present study are available upon reasonable request to the authors

https://zenodo.org/records/14072839

## Supplementary material

**Figure S1:**
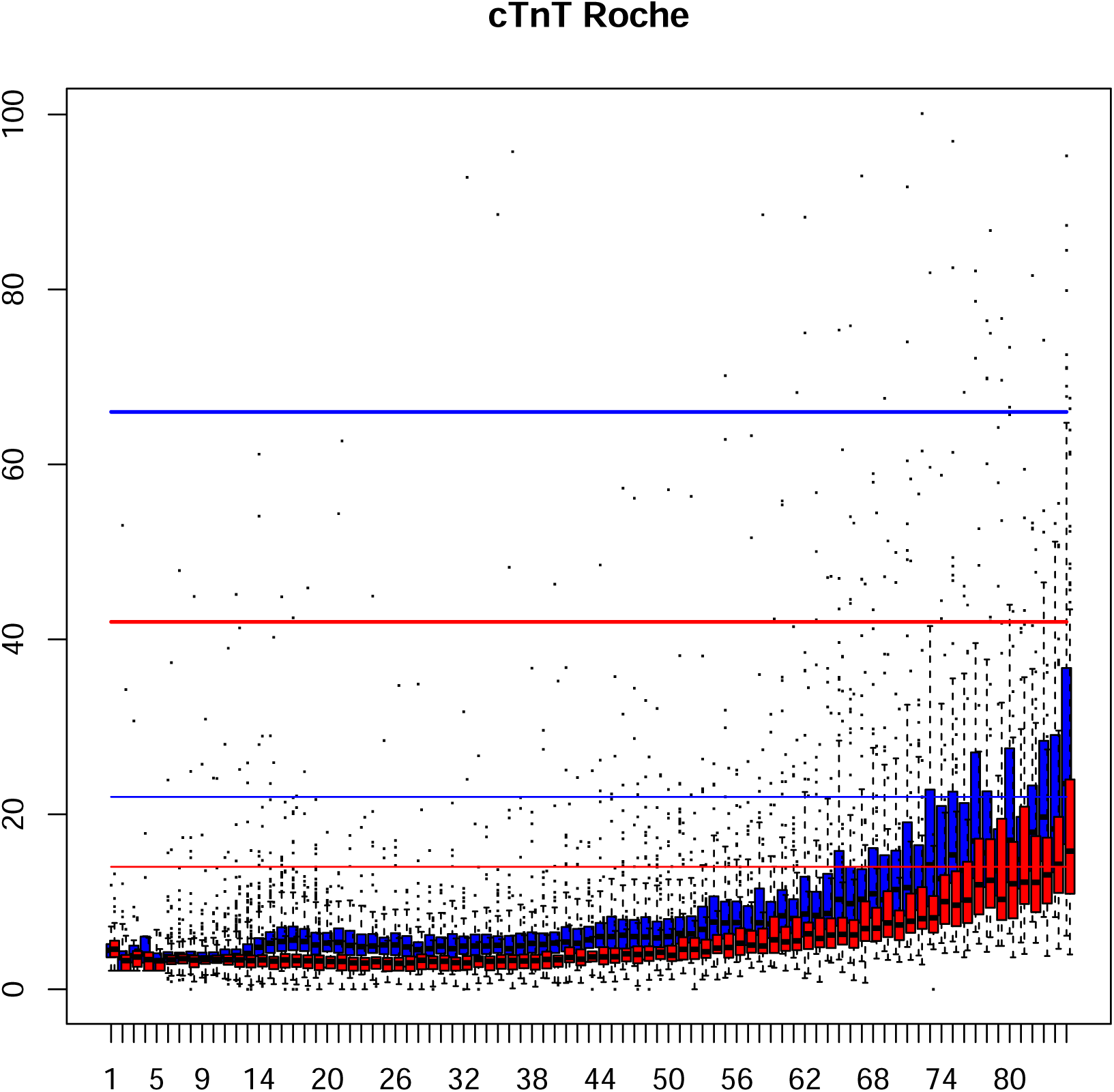
cTnT Roche distribution by age and gender. Red: females. Blue: males. Boxes: median and first and third quartile. Thin horizontal lines: upper limit established by manufacturer. Thick horizontal lines: 3 times the upper limit established by manufacturer.

**Figure S2:**
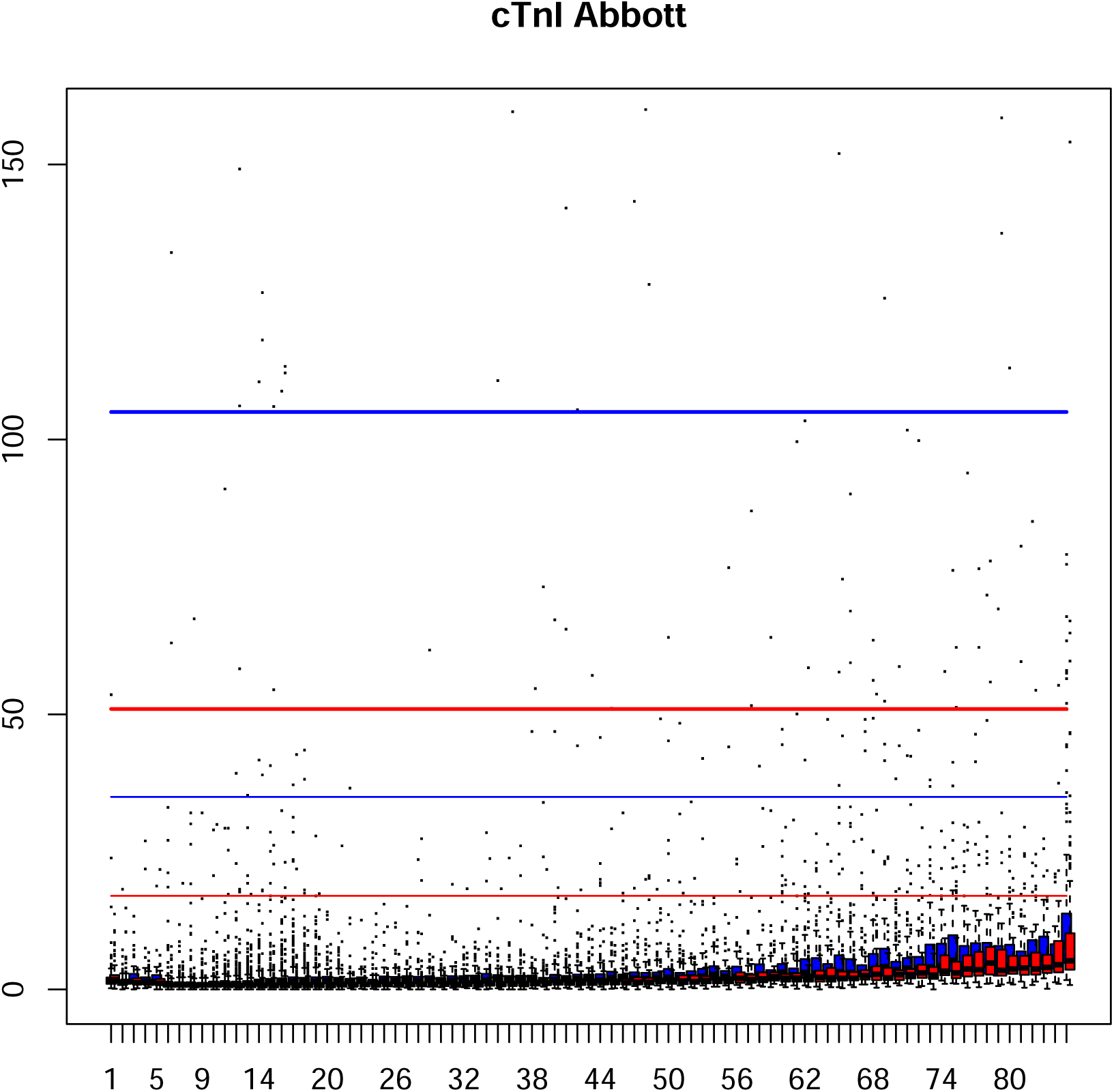
cTnI Abbott distribution by age and gender. Red: females. Blue: males. Boxes: median and first and third quartile. Thin horizontal lines: upper limit established by manufacturer. Thick horizontal lines: 3 times the upper limit established by manufacturer.

**Figure S3:**
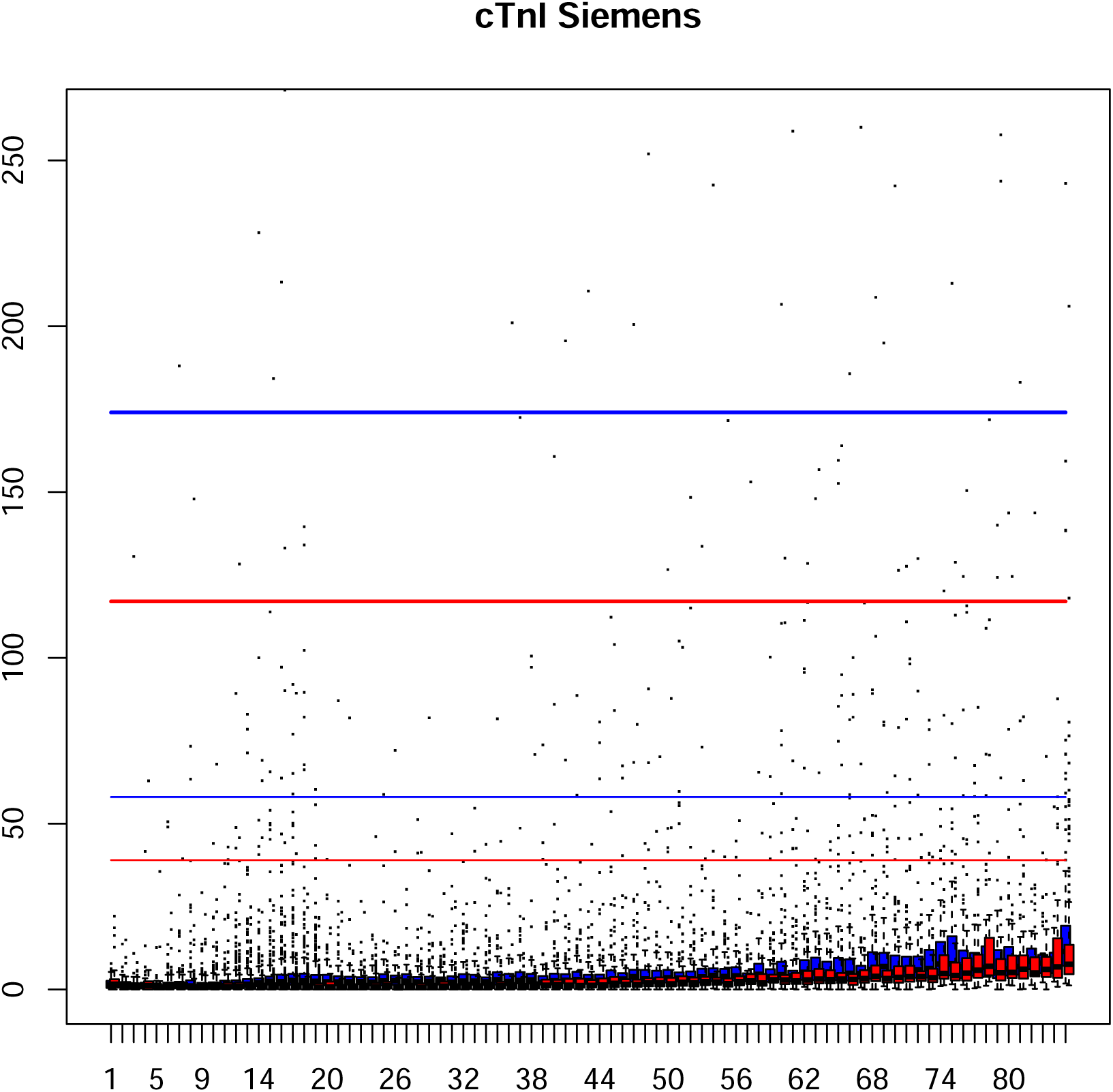
cTnI Siemens distribution by age and gender. Red: females. Blue: males. Boxes: median and first and third quartile. Thin horizontal lines: upper limit established by manufacturer. Thick horizontal lines: 3 times the upper limit established by manufacturer.

**Figure S4:**
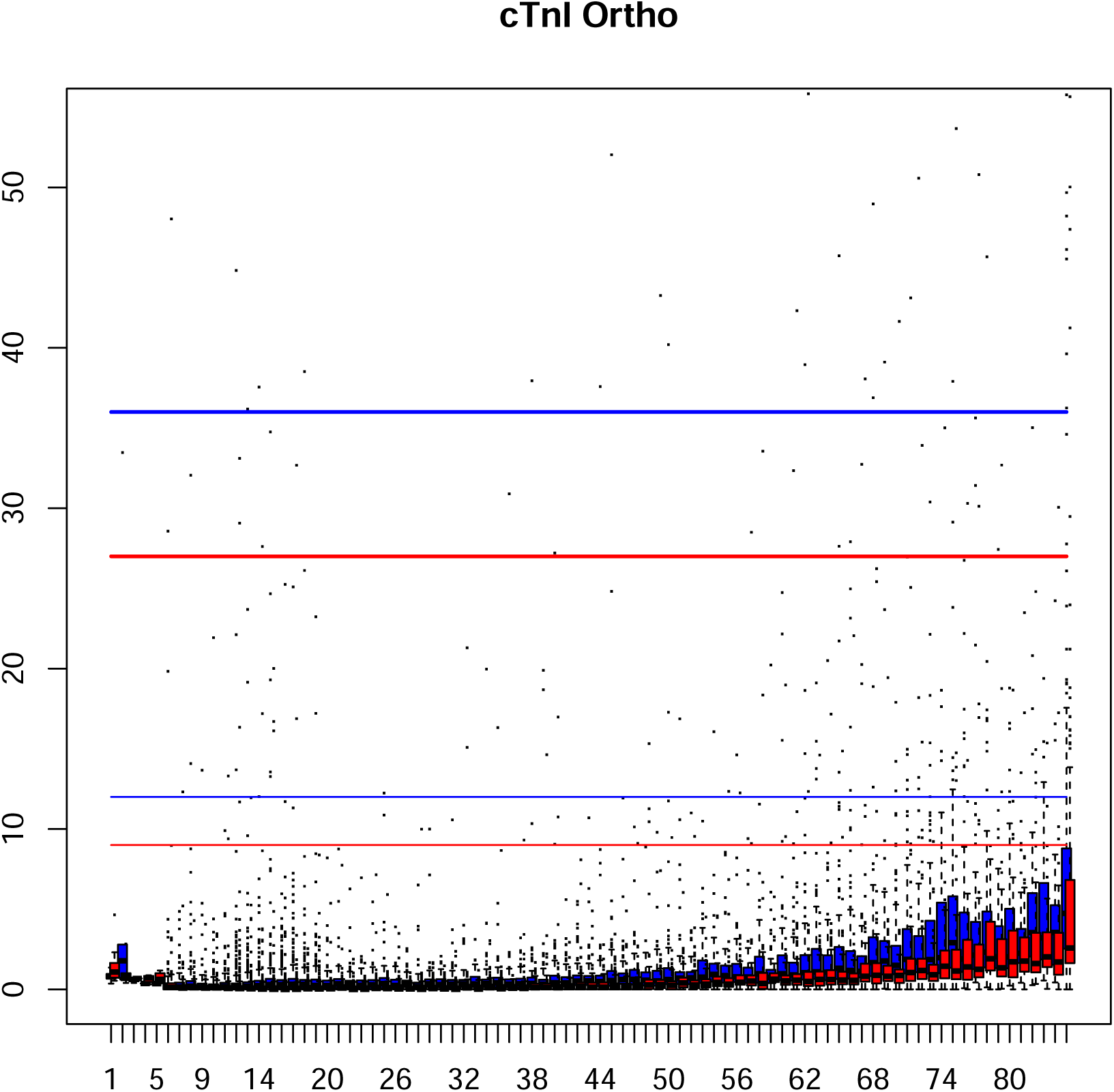
cTnI Ortho distribution by age and gender. Red: females. Blue: males. Boxes: median and first and third quartile. Thin horizontal lines: upper limit established by manufacturer. Thick horizontal lines: 3 times the upper limit established by manufacturer.

**Figure S5:**
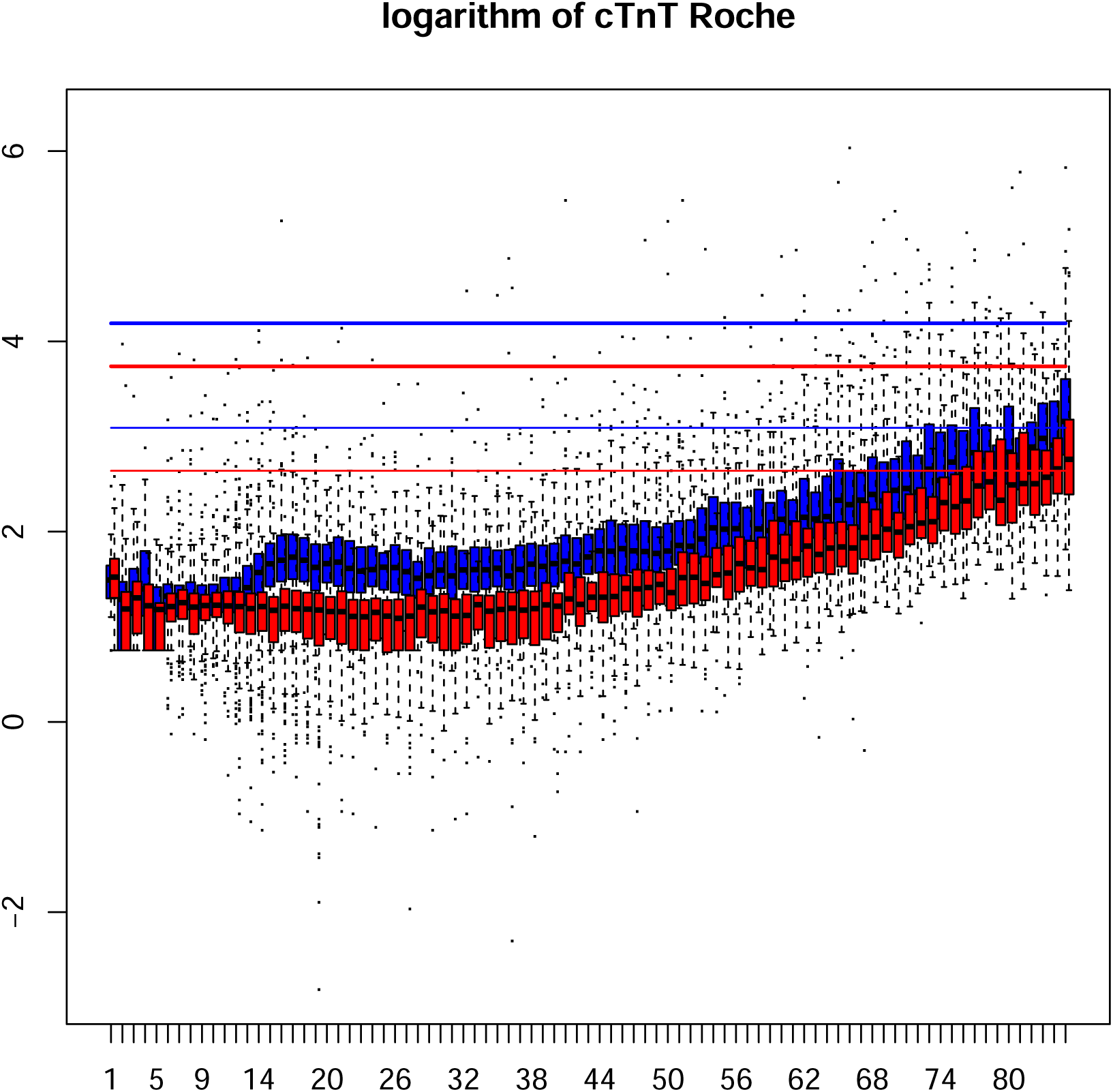
logarithm of cTnT Roche distribution by age and gender. Red: females. Blue: males. Boxes: median and first and third quartile. Thin horizontal lines: upper limit established by manufacturer. Thick horizontal lines: 3 times the upper limit established by manufacturer.

**Figure S6:**
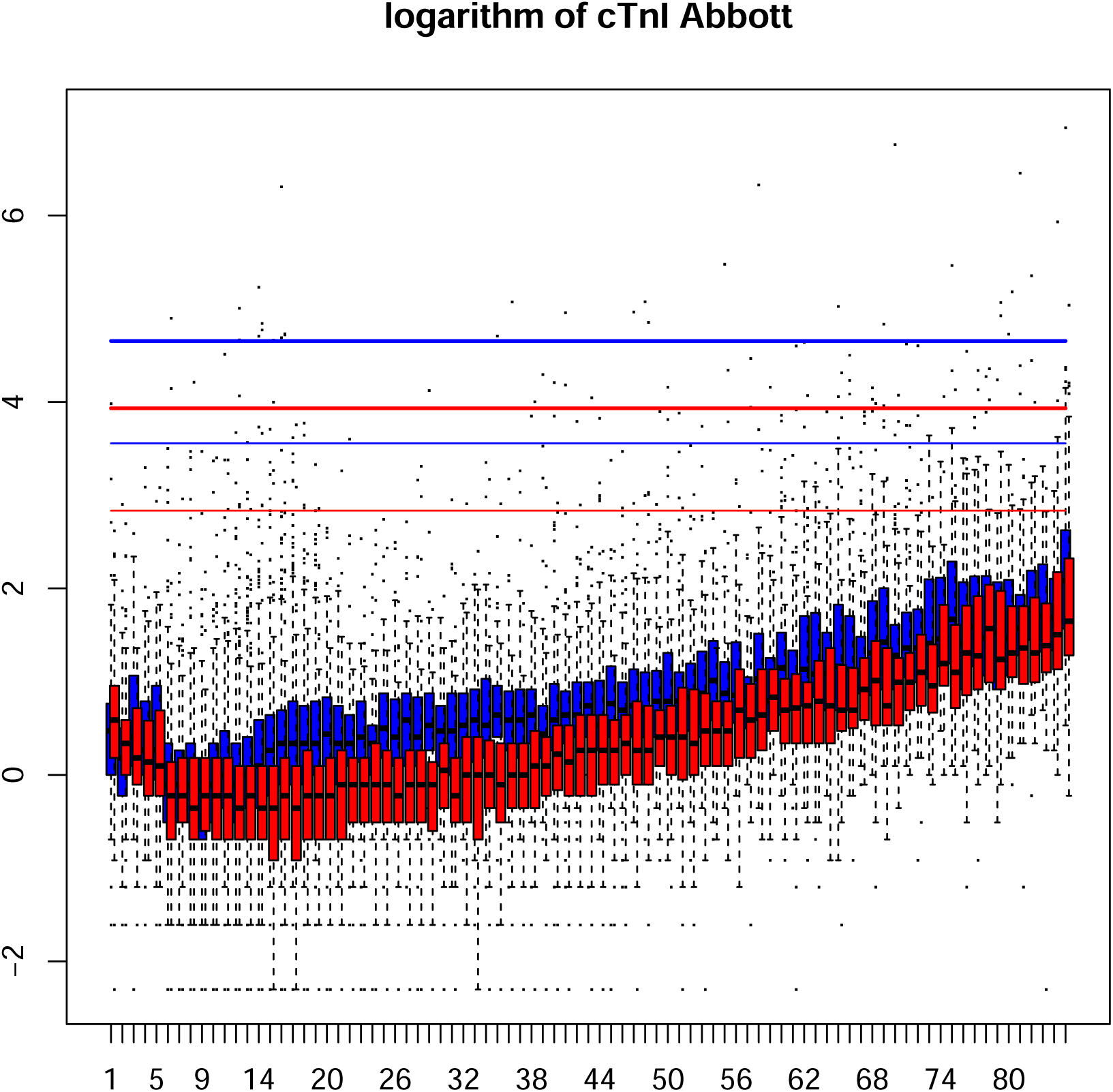
logarithm of cTnI Abbott distribution by age and gender. Red: females. Blue: males. Boxes: median and first and third quartile. Thin horizontal lines: upper limit established by manufacturer. Thick horizontal lines: 3 times the upper limit established by manufacturer.

**Figure S7:**
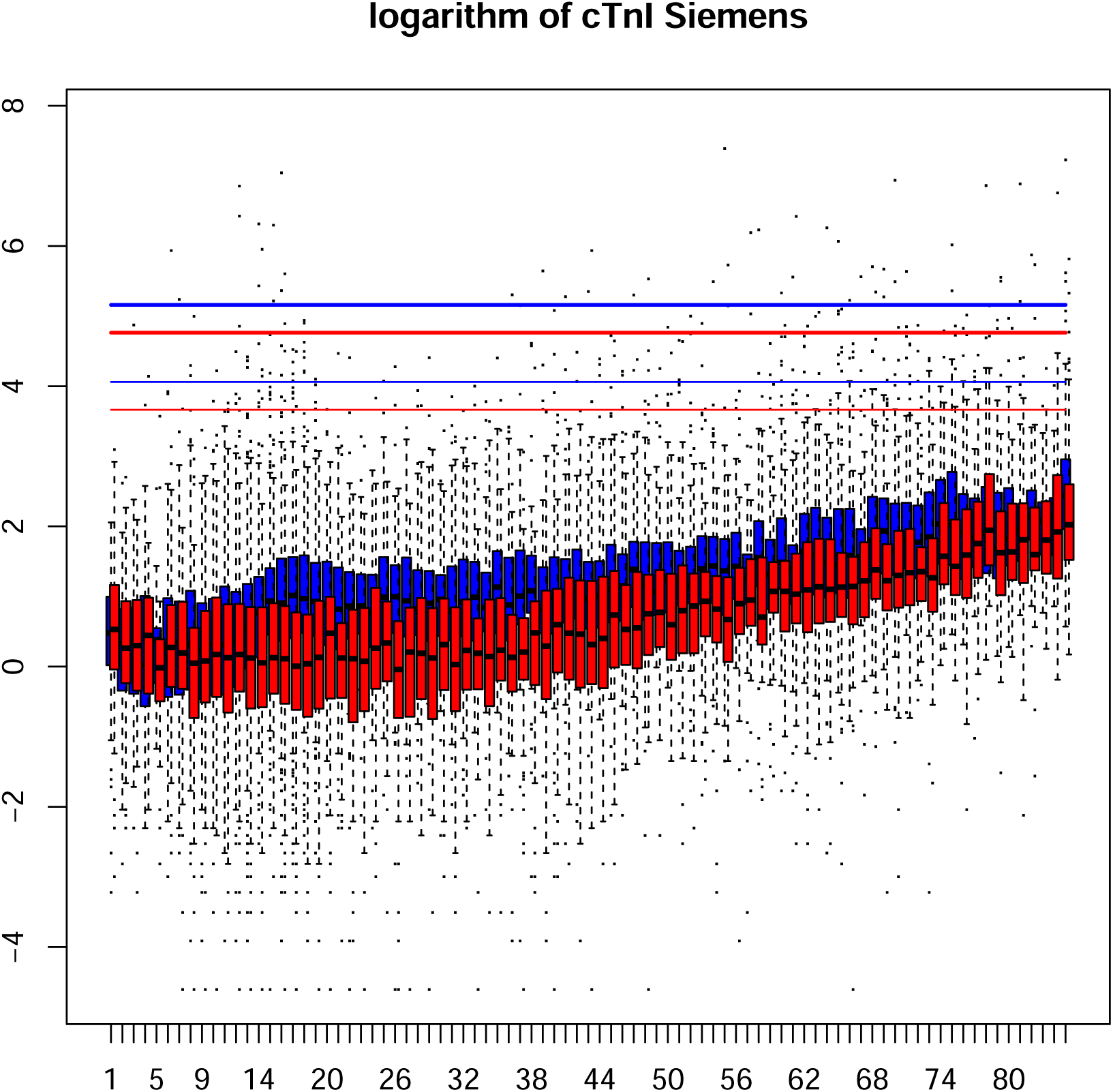
logarithm of cTnI Siemens distribution by age and gender. Red: females. Blue: males. Boxes: median and first and third quartile. Thin horizontal lines: upper limit established by manufacturer. Thick horizontal lines: 3 times the upper limit established by manufacturer.

**Figure S8:**
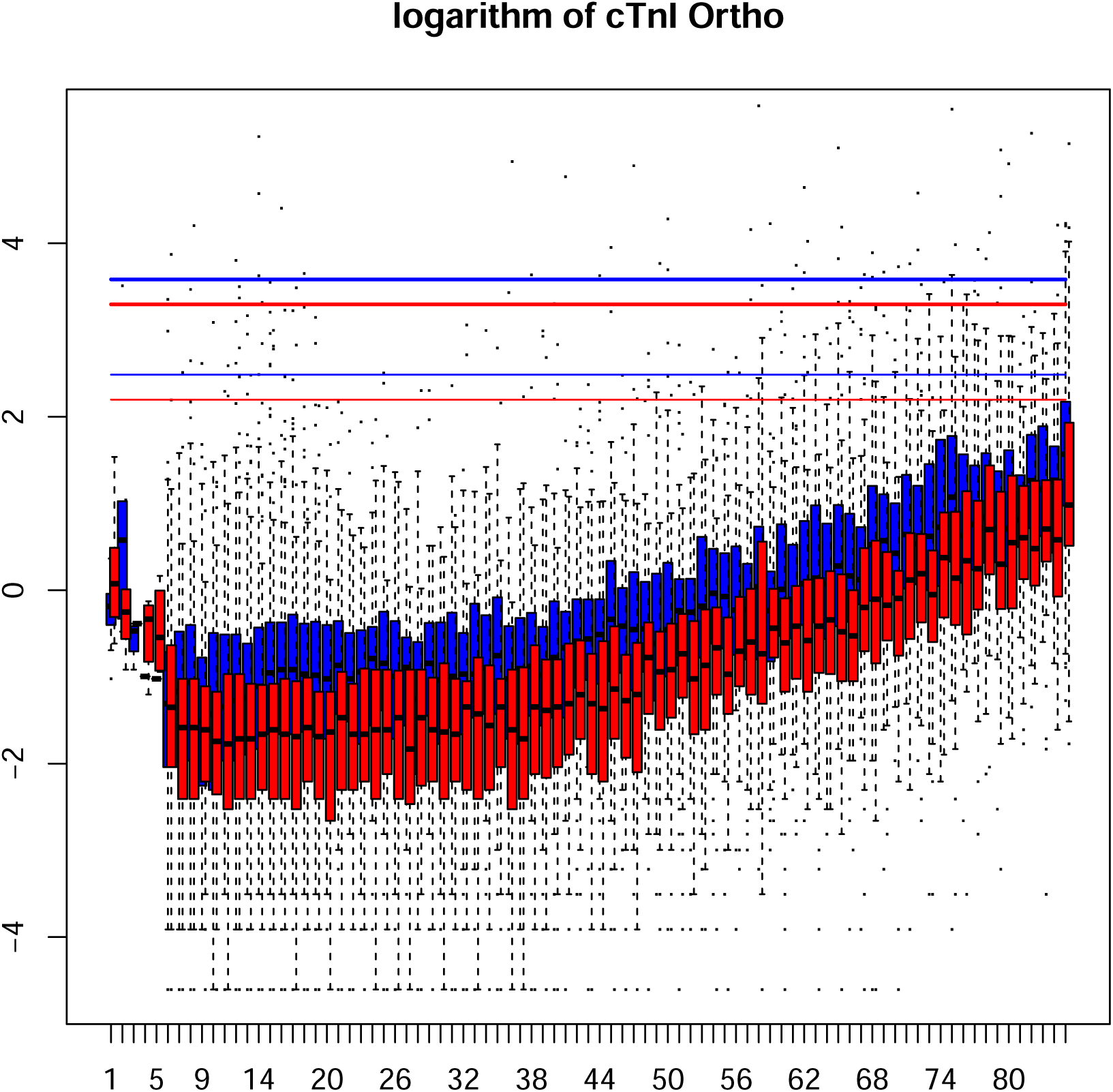
logarithm of cTnI Ortho distribution by age and gender. Red: females. Blue: males. Boxes: median and first and third quartile. Thin horizontal lines: upper limit established by manufacturer. Thick horizontal lines: 3 times the upper limit established by manufacturer.

**Figure S9:**
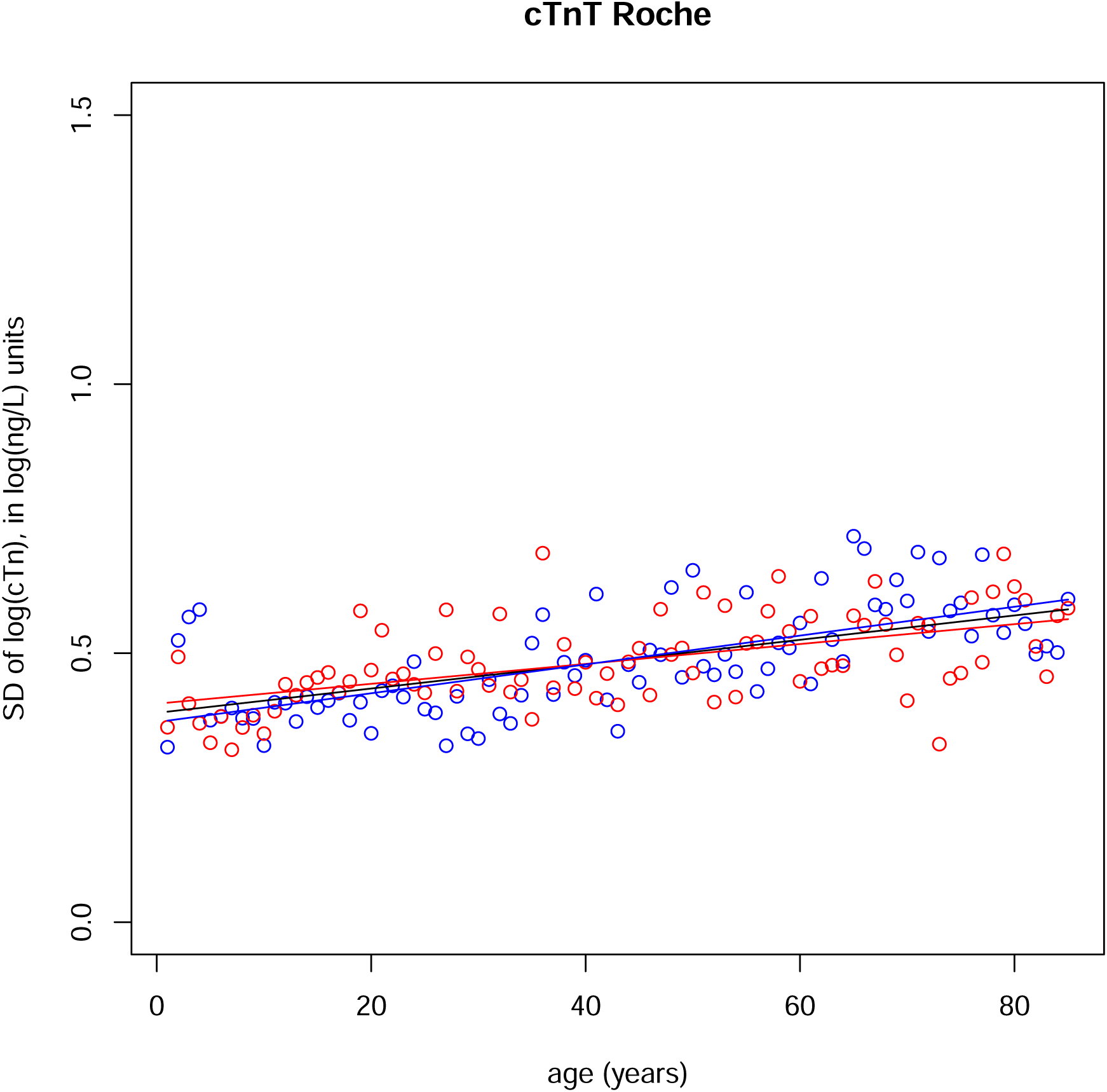
Standard deviations of logarithms of cTnT Roche distributions by age and gender Red: females. Blue: males. Line: linear regression of standard deviations as a function of age, for each gender, black for both males and females.

**Figure S10:**
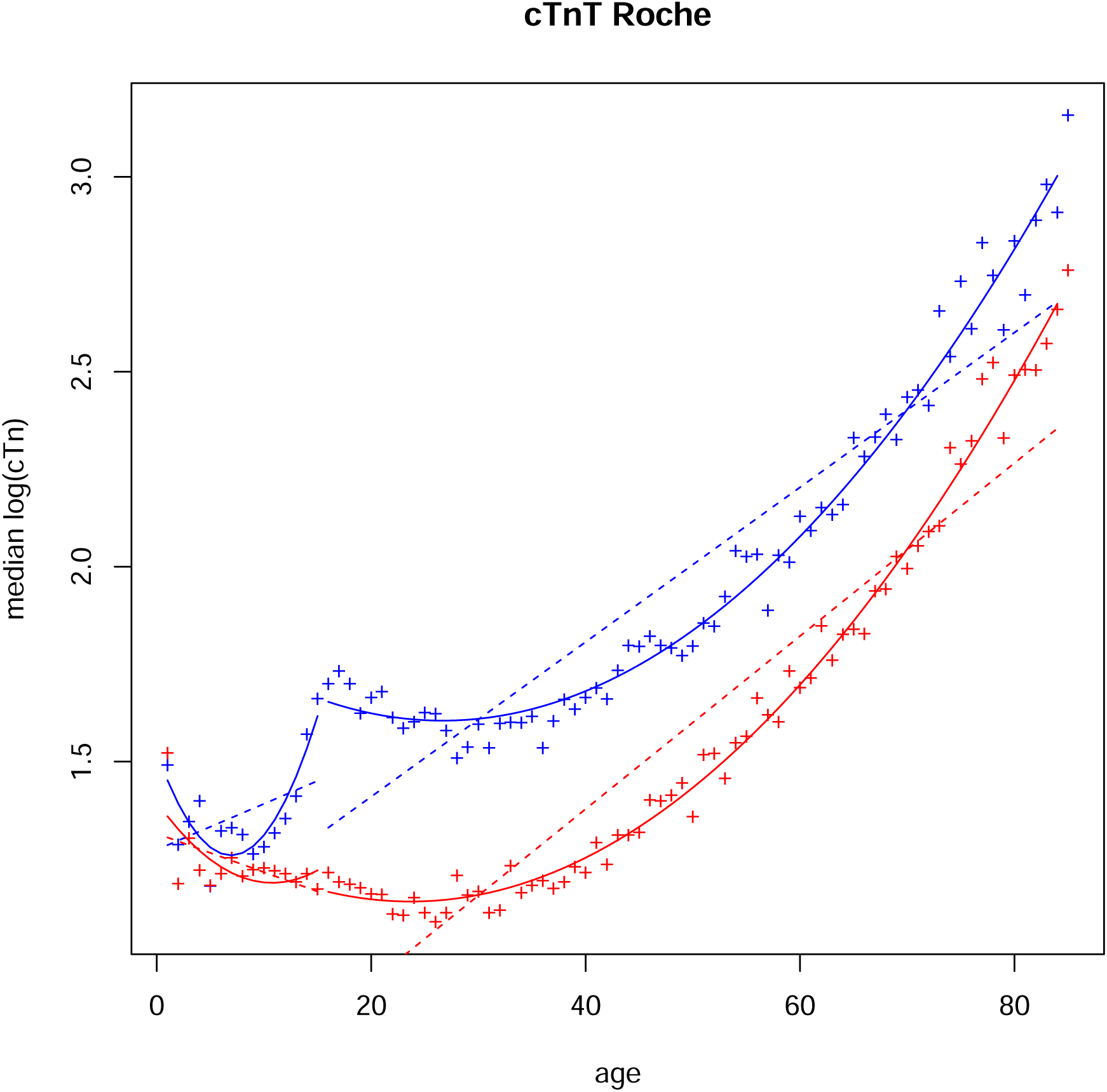
Medians (crosses) of logarithms of cTnT Roche distributions by age and gender Red: females. Blue: males. Dashed lines: linear regression of medians as a function of age for each gender. Curves: quadratic regressions of medians as a function of age for each gender, over1–15 (prepuberal children) and 16–85 age ranges.

**Figure S11:**
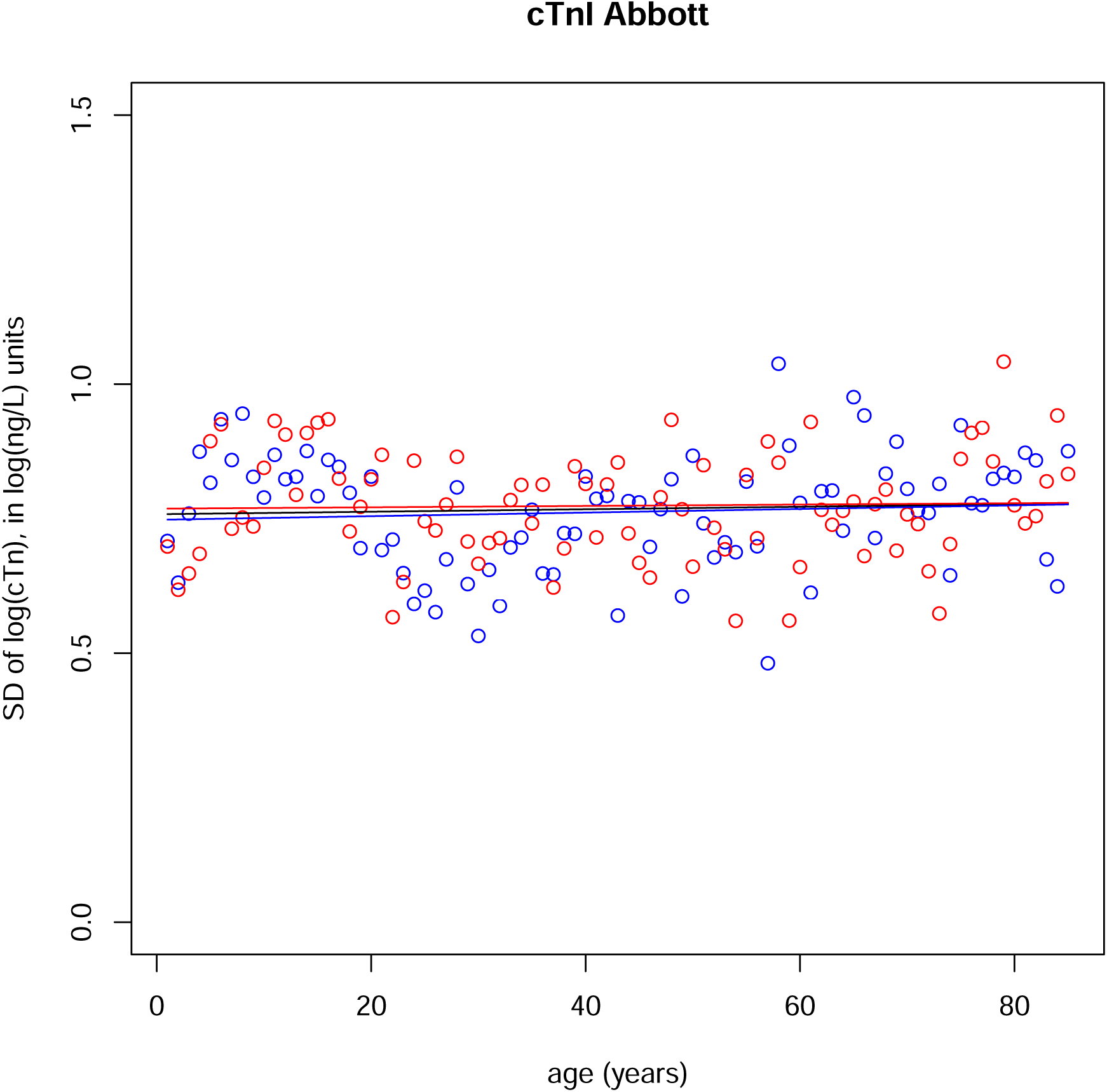
Standard deviations of logarithms of cTnI Abbott distributions by age and gender Red: females. Blue: males. Line: linear regression of standard deviations as a function of age, for each gender, black for both males and females.

**Figure S12:**
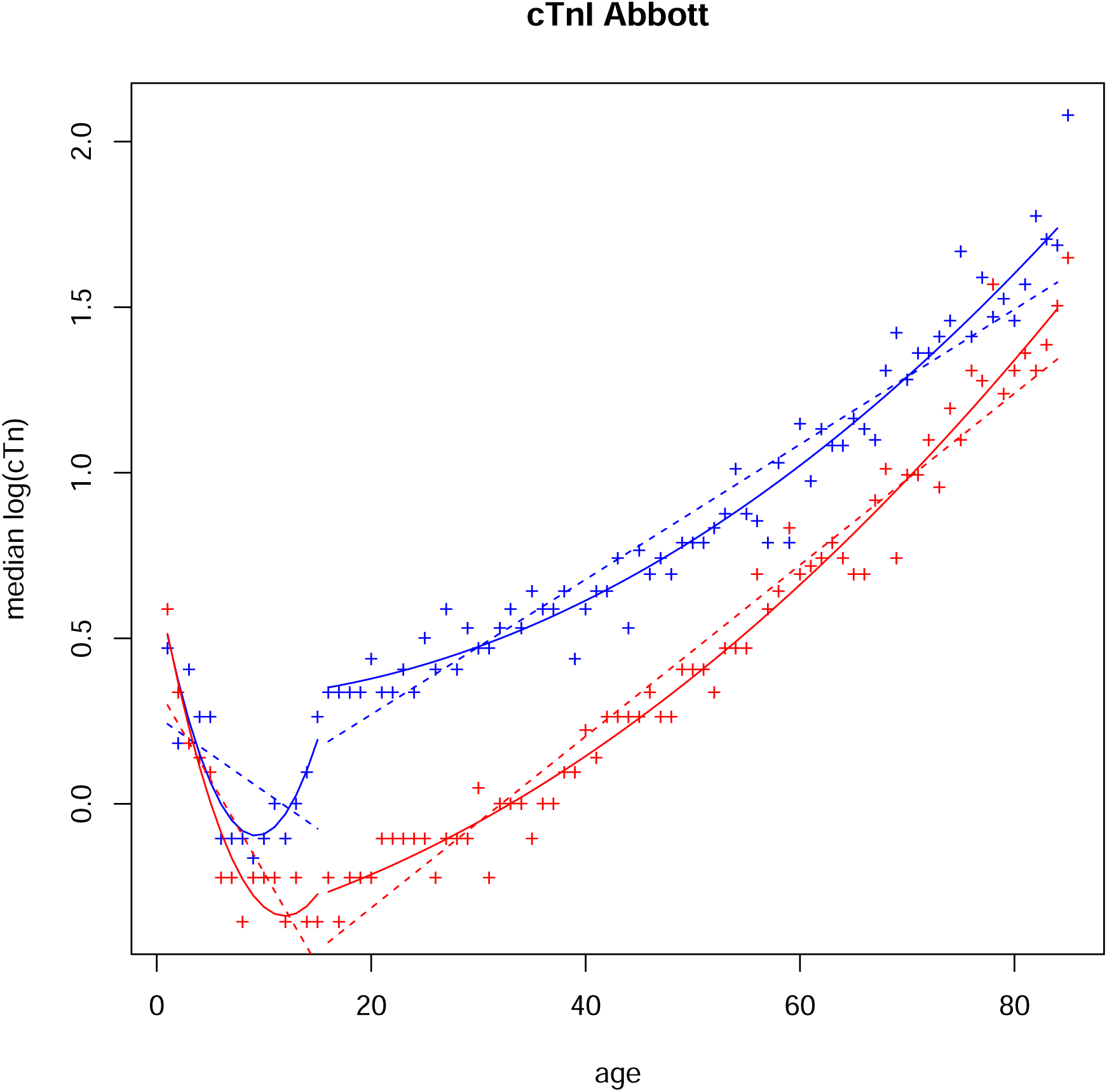
Medians (crosses) of logarithms of cTnI Abbott distributions by age and gender Red: females. Blue: males. Dashed lines: linear regression of medians as a function of age for each gender. Curves: quadratic regressions of medians as a function of age for each gender, over1–15 (prepuberal children) and 16–85 age ranges.

**Figure S13:**
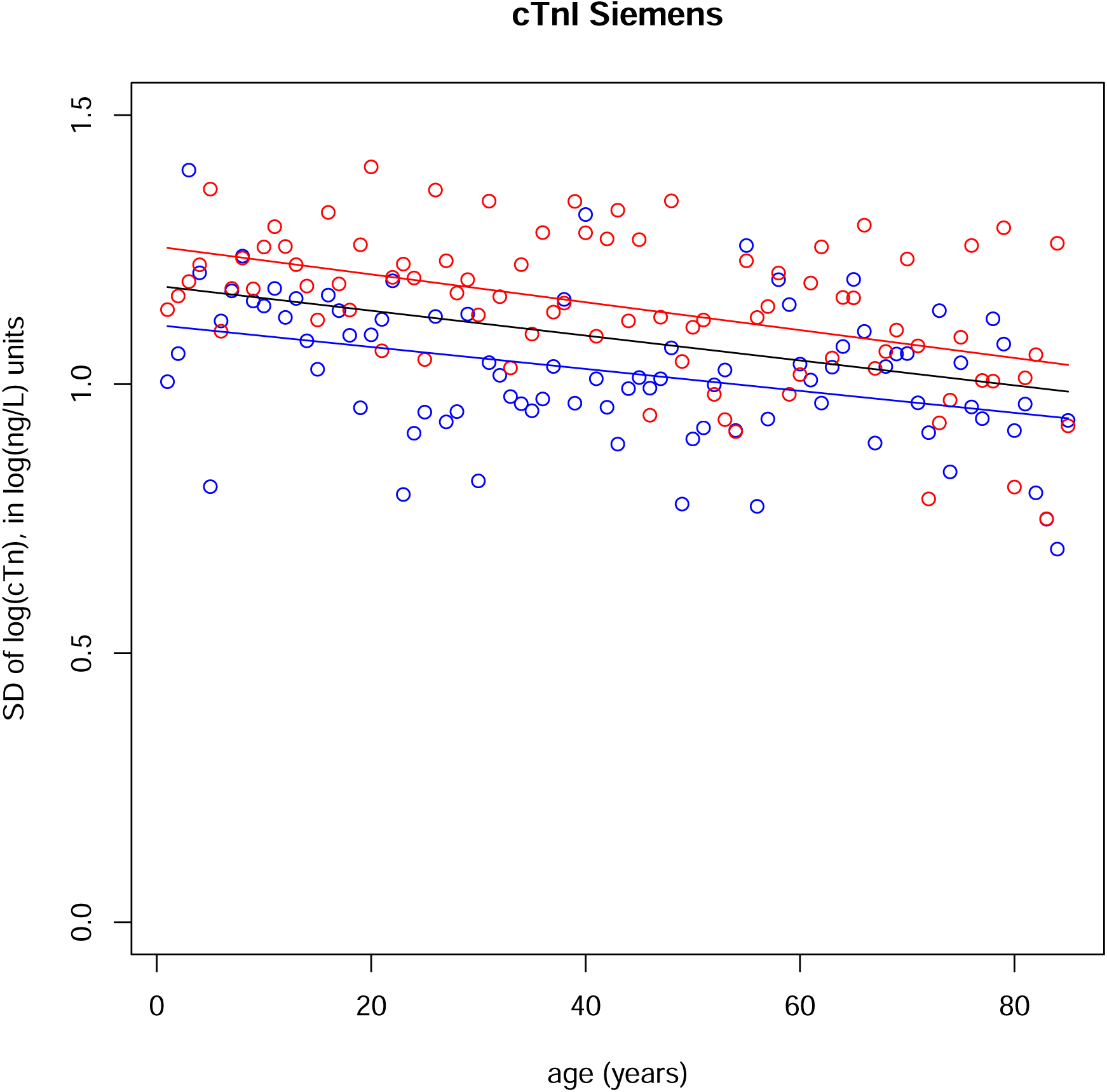
Standard deviations of logarithms of cTnI Siemens distributions by age and gender Red: females. Blue: males. Line: linear regression of standard deviations as a function of age, for each gender, black for both males and females.

**Figure S14:**
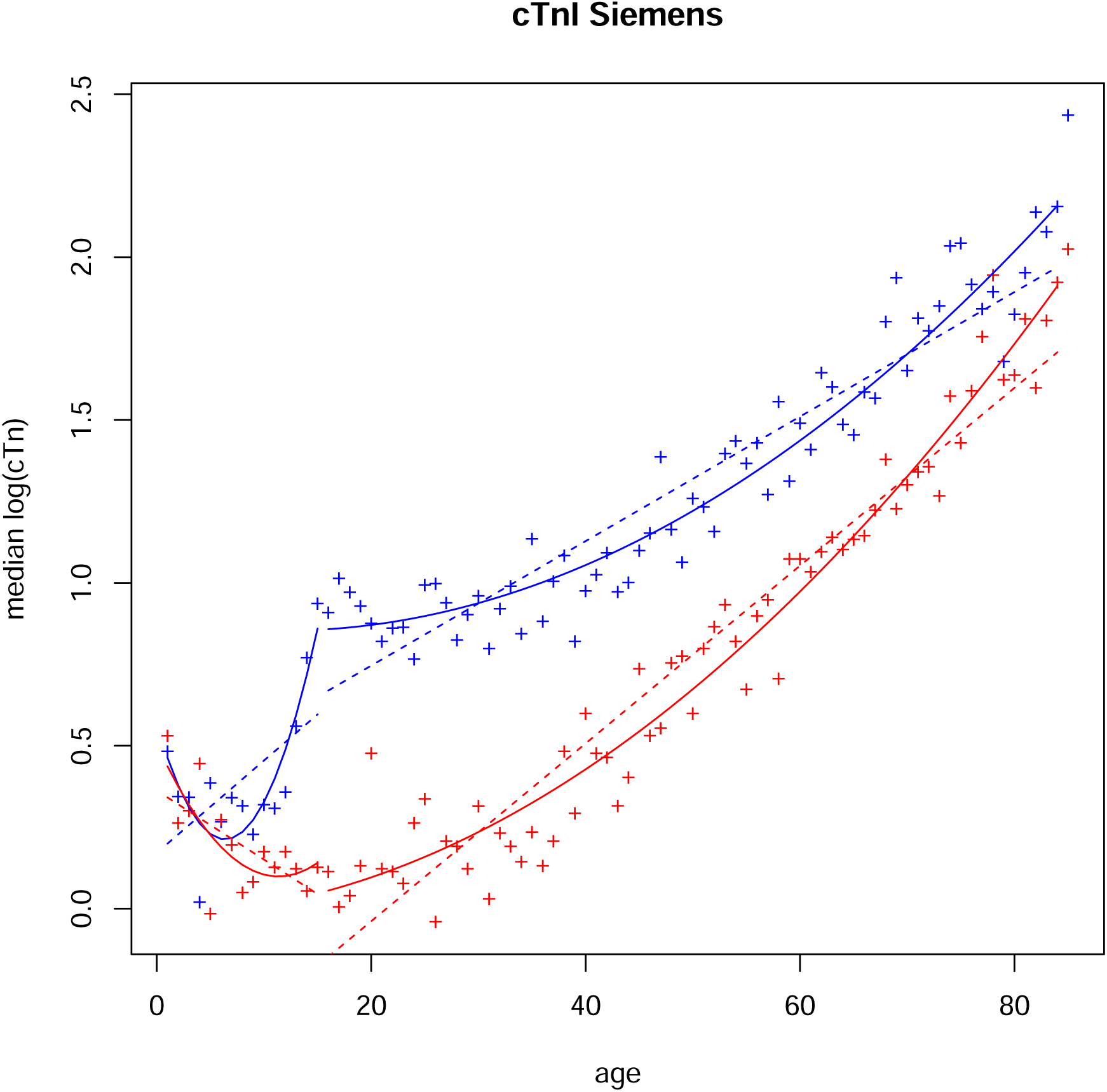
Medians (crosses) of logarithms of cTnI Siemens distributions by age and gender Red: females. Blue: males. Dashed lines: linear regression of medians as a function of age for each gender. Curves: quadratic regressions of medians as a function of age for each gender, over1–15 (prepuberal children) and 16–85 age ranges.

**Figure S15:**
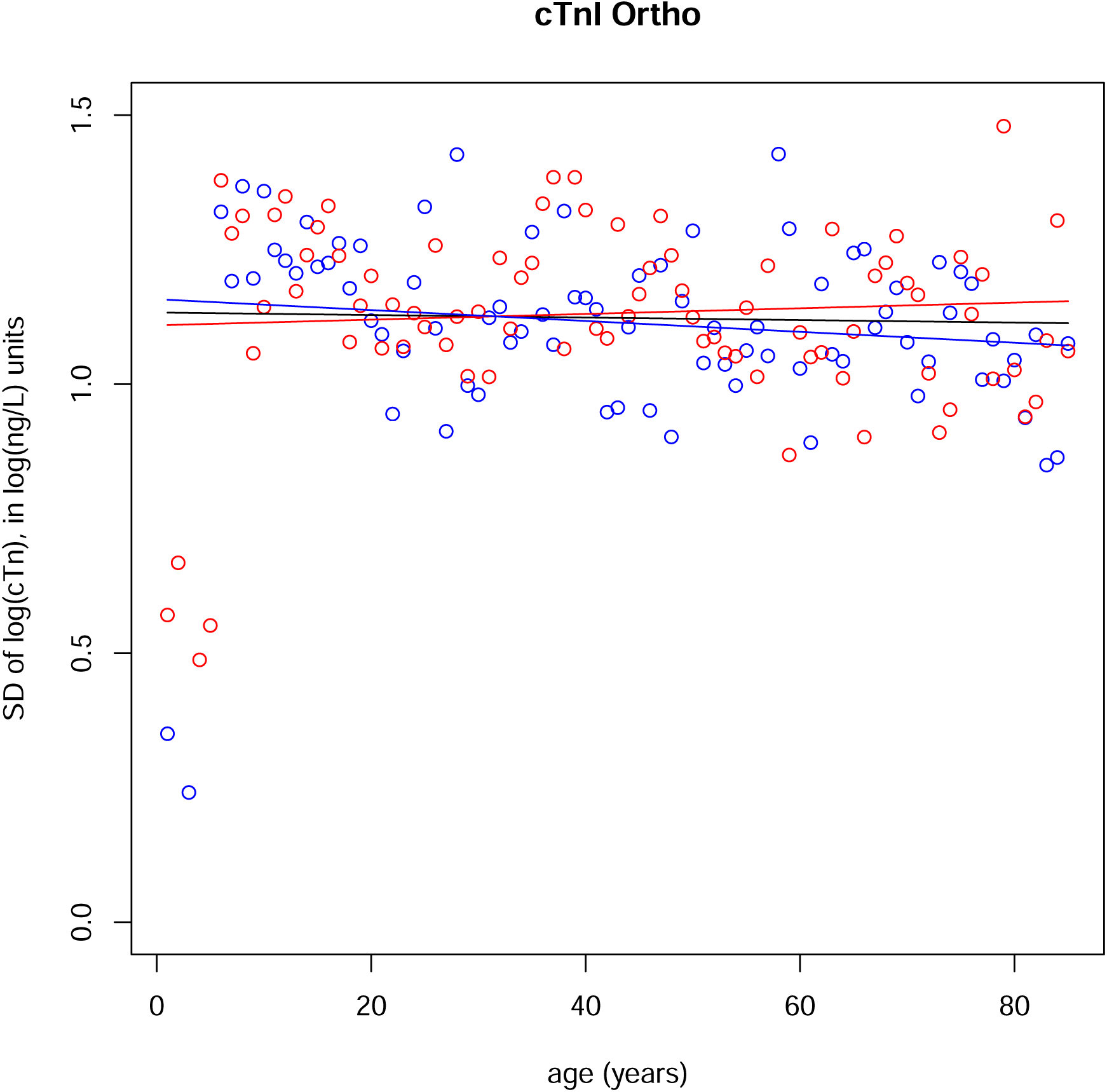
Standard deviations of logarithms of cTnI Ortho distributions by age and gender Red: females. Blue: males. Line: linear regression of standard deviations as a function of age, for each gender, black for both males and females.

**Figure S16:**
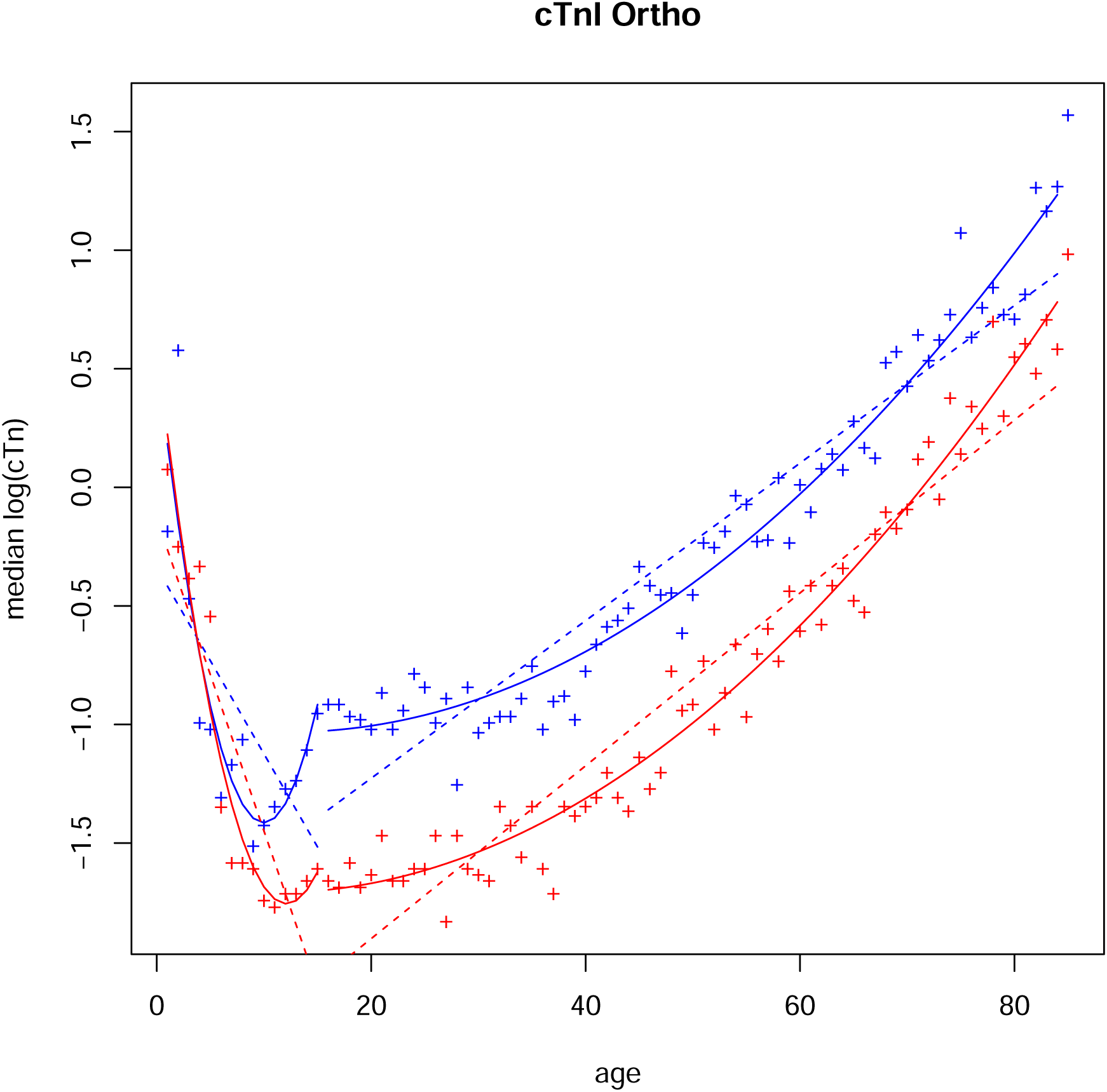
Medians (crosses) of logarithms of cTnI Ortho distributions by age and gender Red: females. Blue: males. Dashed lines: linear regression of medians as a function of age for each gender. Curves: quadratic regressions of medians as a function of age for each gender, over1–15 (prepuberal children) and 16–85 age ranges.

**Figure S17:**
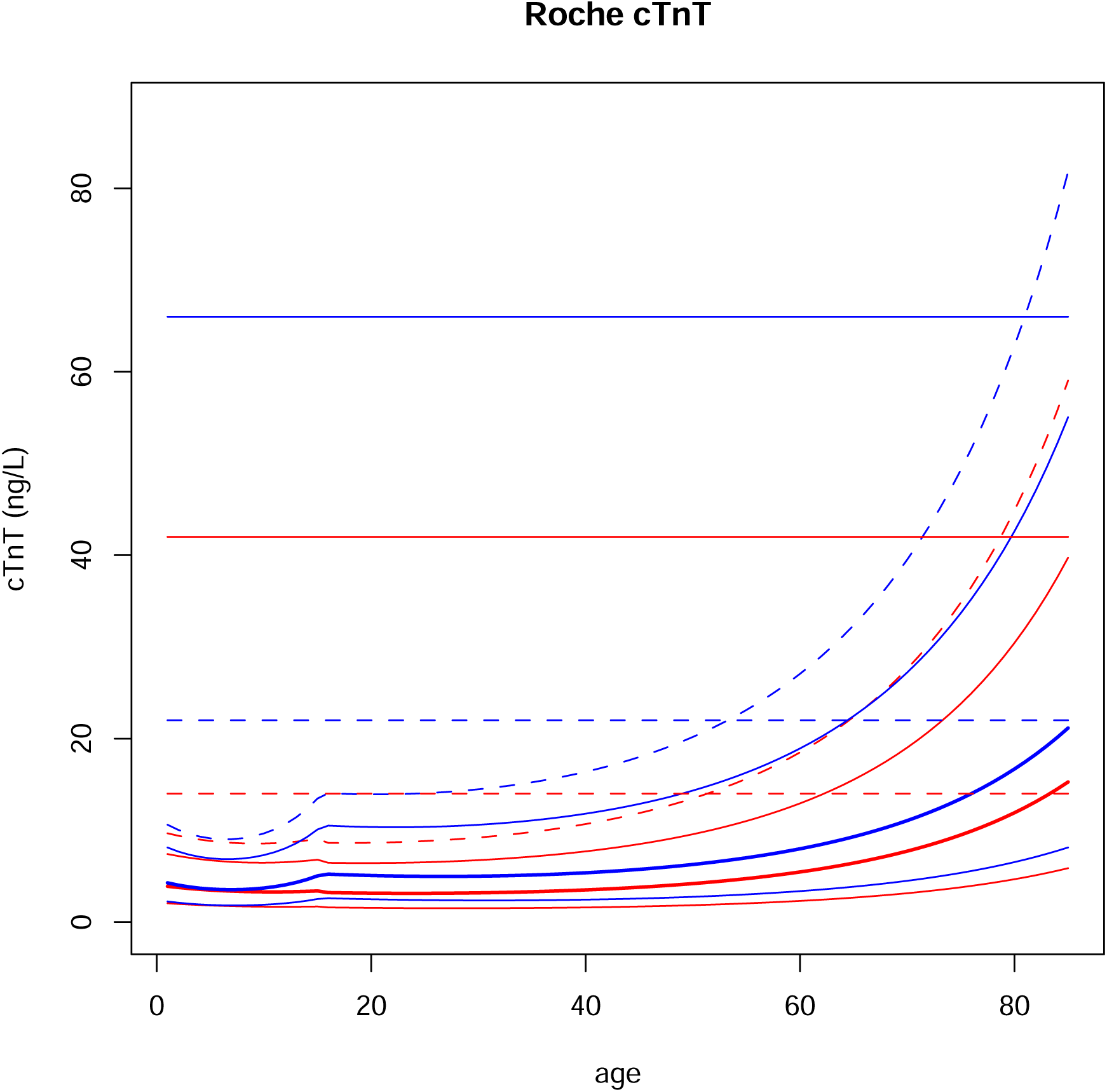
Probability distributions of age and gender dependent models for Roche cTnT. Red: females. Blue: males. Thick curve: median. Thin continuous curves: 0.05 and 0.95 centiles. Dashed curve: 0.99 centile. Dashed horizontal lines: upper limit established by manufacturer. Continuous horizontal lines: 3 times the upper limit established by manufacturer.

**Figure S18:**
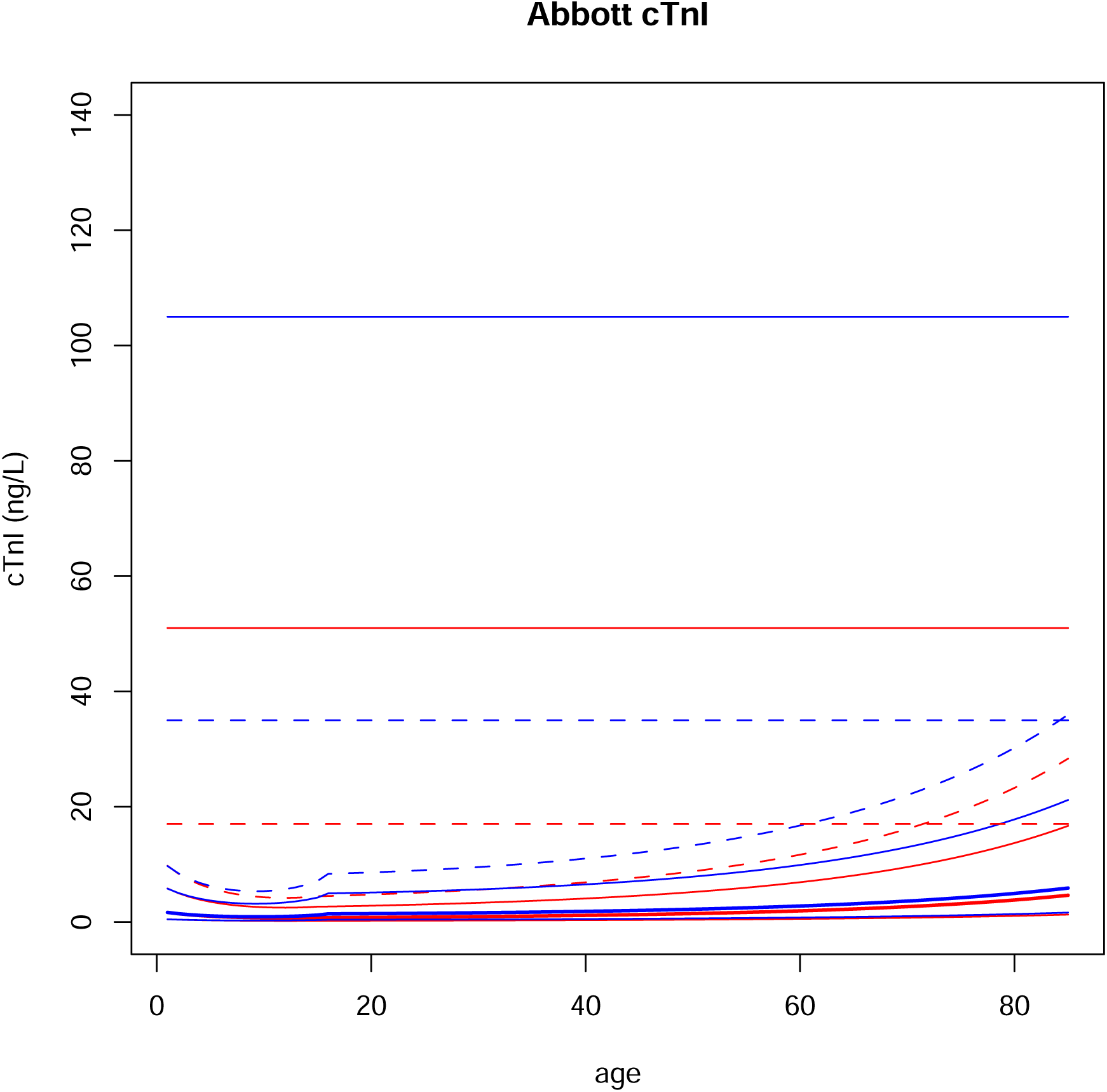
Probability distributions of age and gender dependent models for Abbott cTnI. Red: females. Blue: males. Thick curve: median. Thin continuous curves: 0.05 and 0.95 centiles. Dashed curve: 0.99 centile. Dashed horizontal lines: upper limit established by manufacturer. Continuous horizontal lines: 3 times the upper limit established by manufacturer.

**Figure S19:**
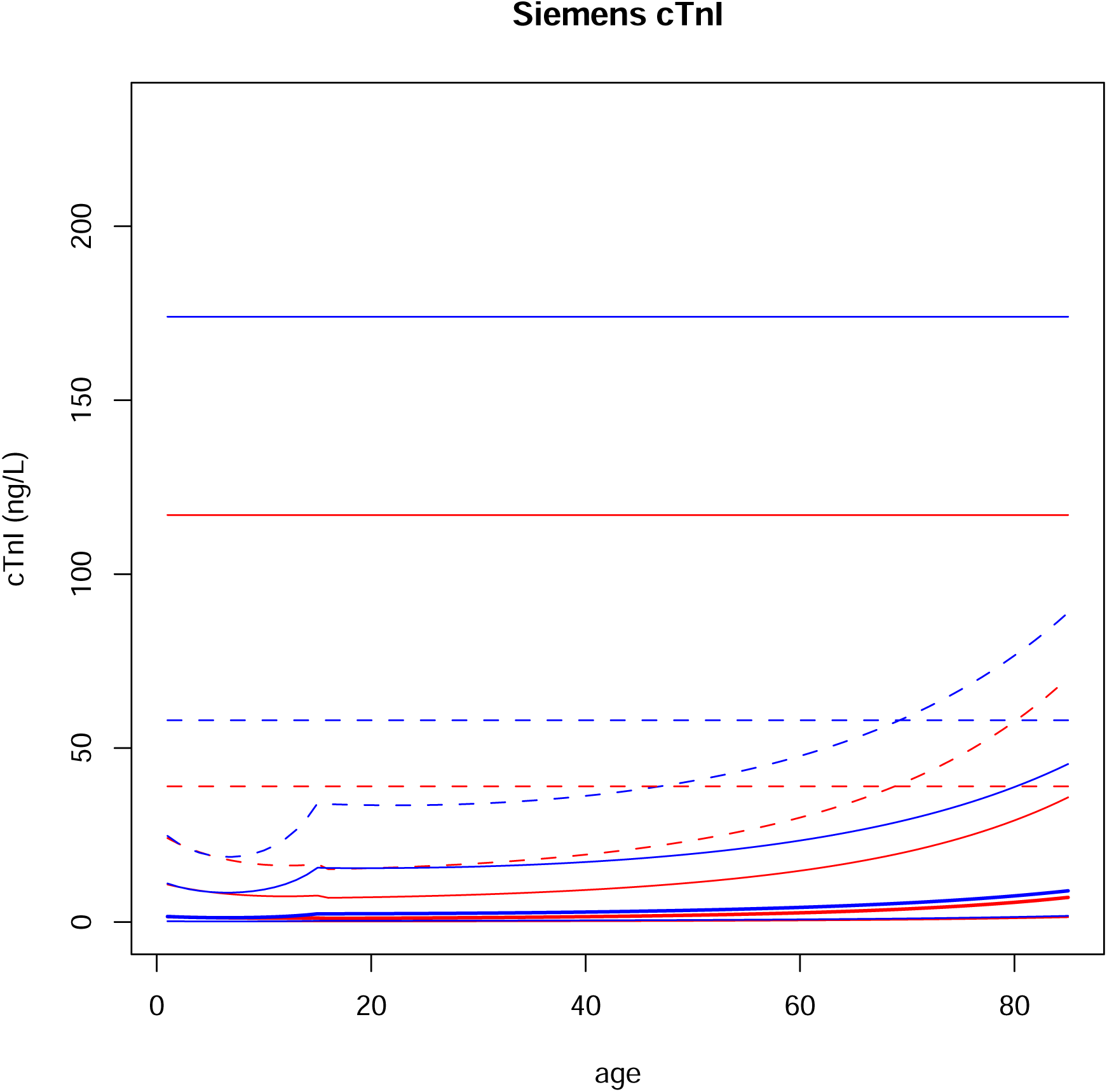
Probability distributions of age and gender dependent models for Siemens cTnI. Red: females. Blue: males. Thick curve: median. Thin continuous curves: 0.05 and 0.95 centiles. Dashed curve: 0.99 centile. Dashed horizontal lines: upper limit established by manufacturer. Continuous horizontal lines: 3 times the upper limit established by manufacturer.

**Figure S20:**
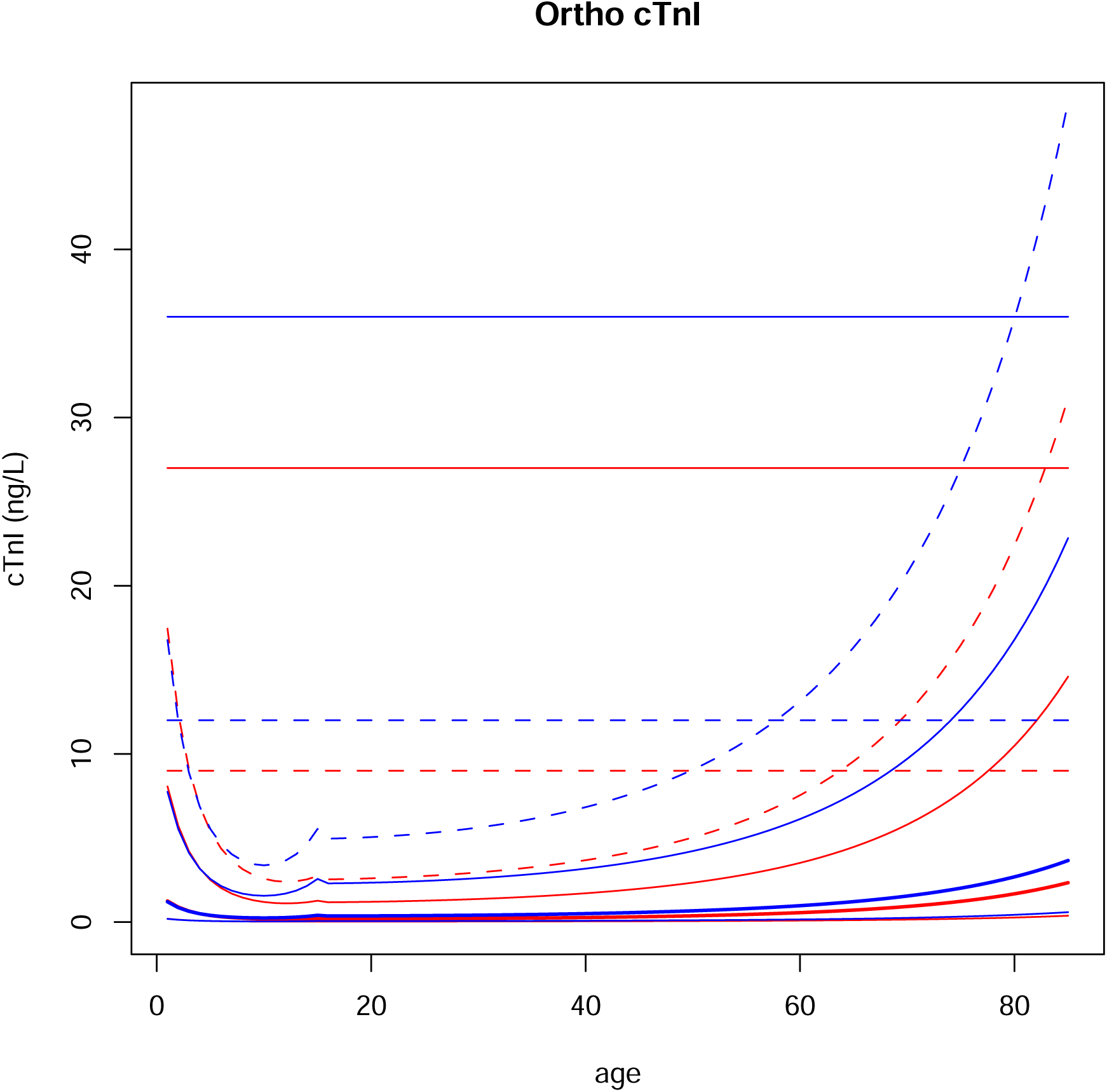
Probability distributions of age and gender dependent models for Ortho cTnI. Red: females. Blue: males. Thick curve: median. Thin continuous curves: 0.05 and 0.95 centiles. Dashed curve: 0.99 centile. Dashed horizontal lines: upper limit established by manufacturer. Continuous horizontal lines: 3 times the upper limit established by manufacturer.

**Table S1:**
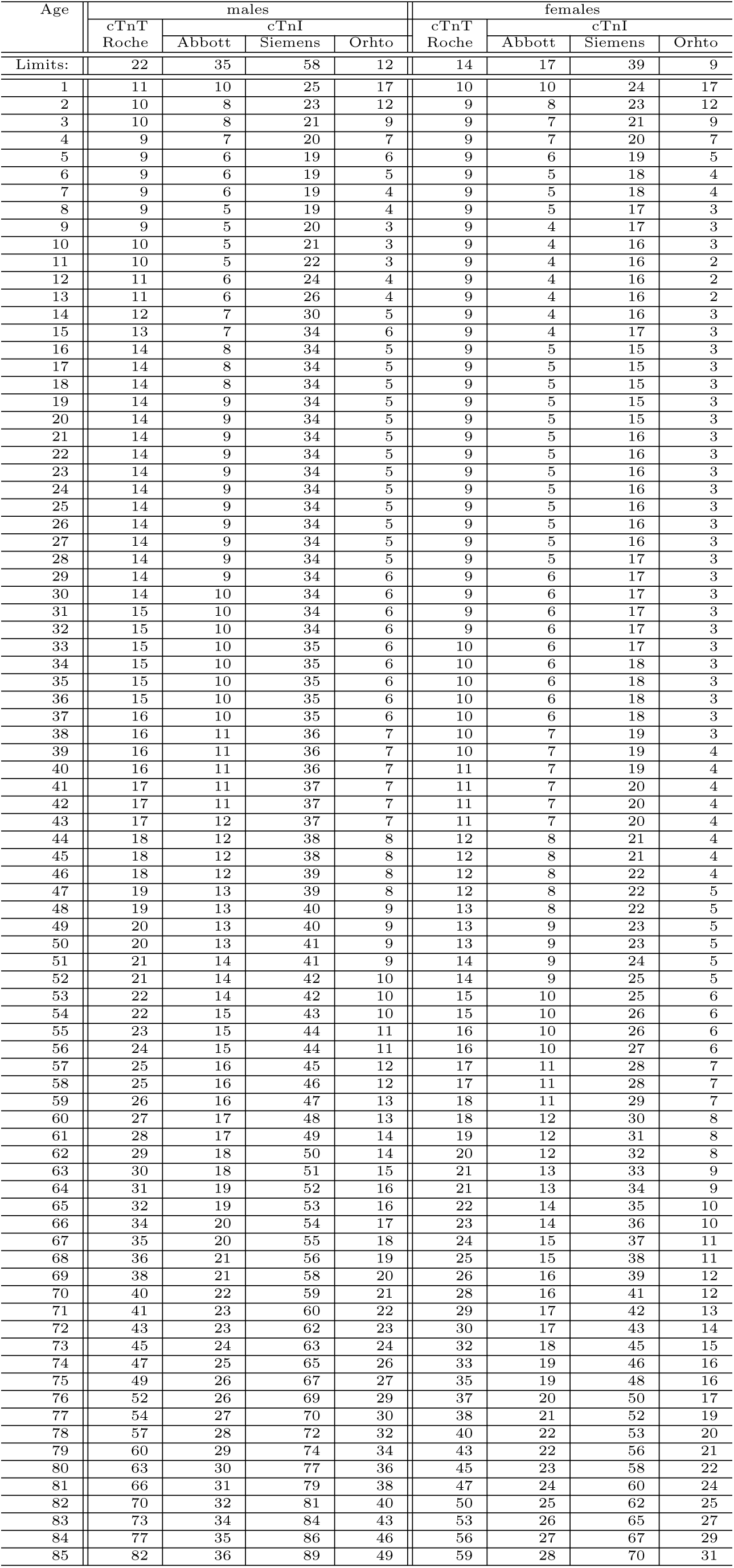
General population limits for the 99-th percentile.

**Table S2:**
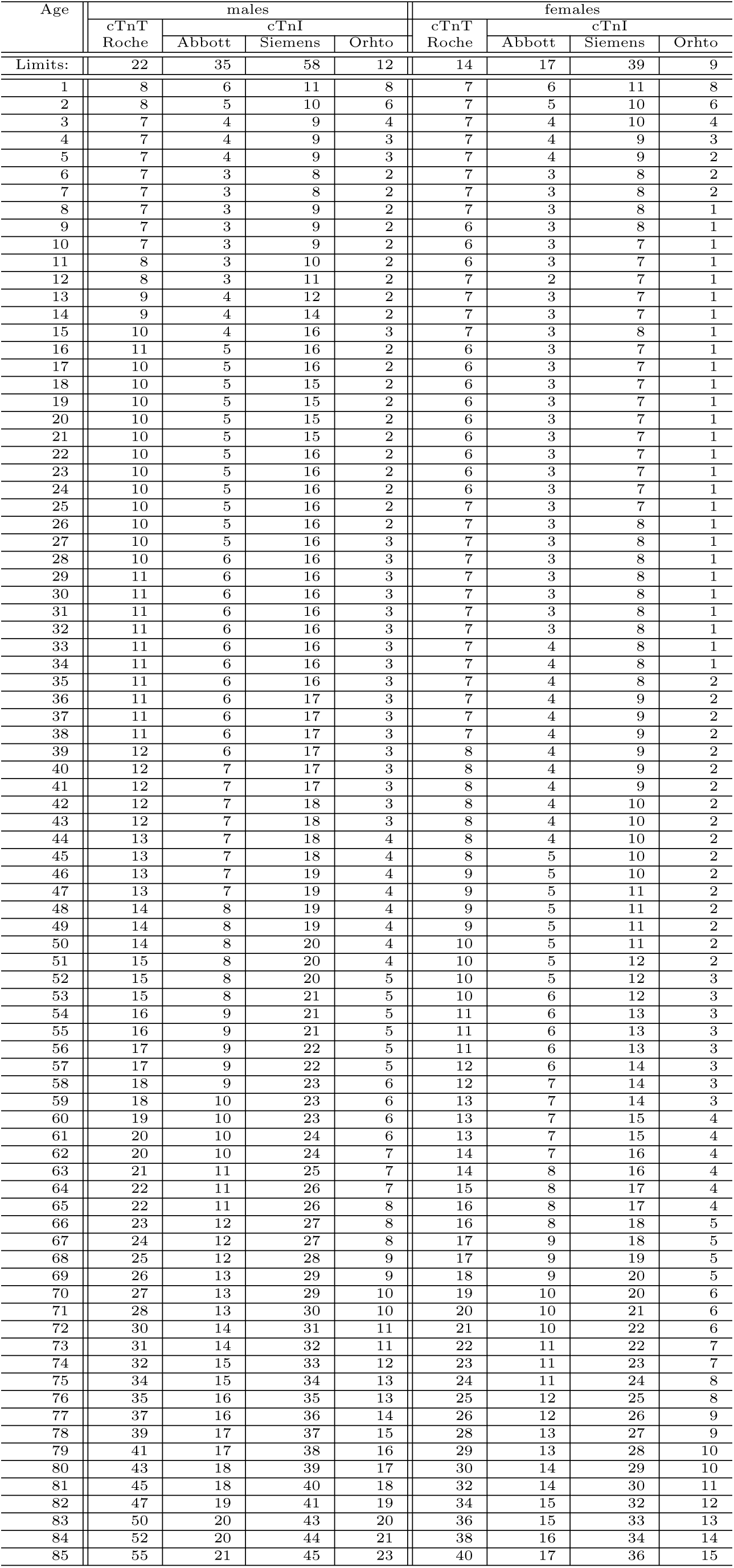
General population limits for the 95th percentile.

**Table S3:**
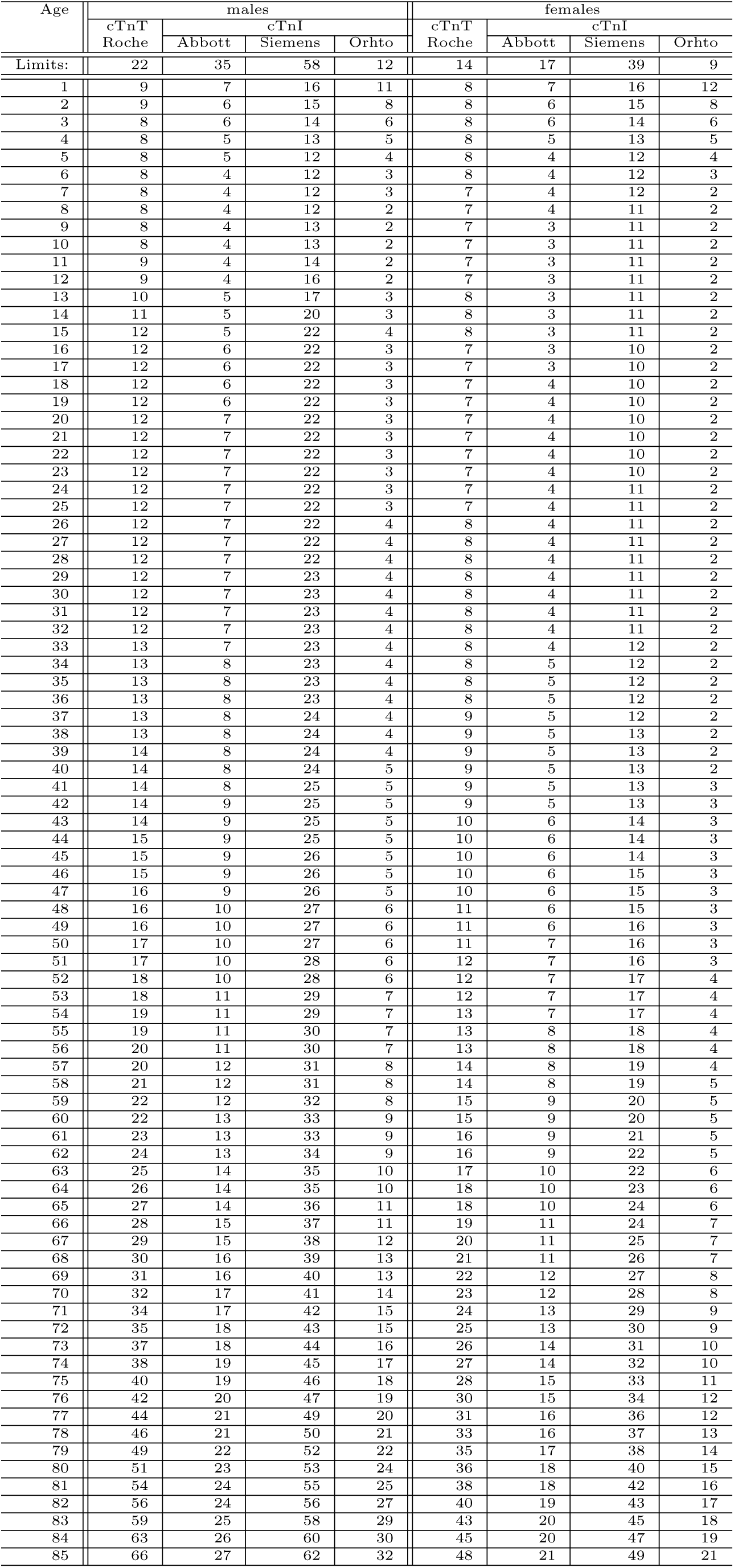
General population limits for the 97.5th percentile.

